# Management of splenic injury in children and young adults: a survey of surgeons investigating factors responsible for variations in care using the Theoretical Domains Framework

**DOI:** 10.1101/2023.07.26.23293058

**Authors:** Susan E Adams, Andrew JA Holland, Julie Brown

## Abstract

Objective: To identify differences in practice and behavioural drivers in adult and paediatric surgeons related to the management of splenic injury in children and young people.

Background: Despite the existence of guidelines, there are variations in the care of children with splenic injuries. There are no specific guidelines for the care of young adults, who may be managed according to either paediatric or adult practices. The drivers of variation in trauma management between adult and paediatric surgeons have not been examined through an implementation science lens.

Methods: The COM-B model of behaviour and theoretical domains framework were used to construct a survey which was delivered to a cross-section of adult general surgeons and paediatric surgeons working in public hospitals throughout NSW, Australia. The capability, opportunity, and motivation for management decisions were analysed. Outcome variables were compared between the practitioner groups. Statistical significance was set at p < 0.05.

Results: Eighty (26.4%) responses met the inclusion criteria. Significant differences between adult and paediatric surgeons were identified in terms of: capability (surgeon training and experience); opportunity (hospital, personnel, and resources); and motivation (comfort with splenic injury care at different ages). In managing splenic injury, pediatric surgeons tended to follow pediatric guidelines while adult surgeons followed adult guidelines, making some adjustments for age. All agreed guidelines had the potential to improve care.

Conclusions: This study identified several behavioural drivers for observed variations in the care of splenic injury in children and young adults. The results indicate that contextually relevant guidelines for managing splenic injury in children and young people across any setting may be needed to reduce disparities in care. These should be underpinned by interventions designed to further address the drivers of surgeon behaviour to optimise uptake. Furthermore, splenic injury care is a clinical indicator of the quality of trauma care more broadly. Addressing variation in its care has the potential to translate to other trauma system improvements.

**HIGHLIGHTS:** Unwarranted variations in splenic injury management have been reported.

This is the first examination of surgeon behaviour in the provision of trauma care through an implementation science lens.

Differences in the capability, opportunity, and motivation of surgeons in providing evidence-based care to children and young adults with splenic injury were found.

Contextually relevant guidelines, underpinned by supporting interventions addressing these behavioural drivers, are needed to reduce disparities in care.

## INTRODUCTION

The spleen is the most commonly injured abdominal organ at all ages. (1) In pediatric guidelines, non-operative management is the accepted standard of care,(2) with almost 100% of isolated splenic injury cases able to be successfully managed in this way.(3, 4) This has major benefits, including avoiding infective risks associated with asplenia (5) and complications of laparotomy and angioembolisation (6, 7). In adults, the concept of selective non-operative management has emerged, and guidelines for adult splenic trauma management have also been developed and updated.(8)

Despite the existence of guidelines for many conditions, unwarranted variations in healthcare are well described.(9, 10) This is evident in the care of children and young adults with splenic injuries: In the United Kingdom, North America, and Australia, hospital type has been correlated with intervention, including operation and angioembolisation.(3, 11–19) It is unknown what factors may be influencing management decisions in these different settings and age groups.

There is a need to identify effective mechanisms to improve the quality of trauma management and reduce variations in care. Splenic injury management is a quality indicator of abdominal trauma management in trauma systems more broadly,(3) so identifying effective ways to optimise its care may provide important lessons for optimised trauma management universally. Previous work indicates a wide variety of patient, hospital, and surgeon factors that may influence management decisions.(13, 18) However, there has been no work examining the drivers of variation in trauma management between adult and pediatric surgeons through an implementation science (IS) lens.

There is growing acknowledgement that grounding clinical research in the principles of IS is critical to the translation of research outcomes to produce sustainable improvement of health care practices such as surgical care.(20, 21)

In our previous work, we have undertaken the first IS step by identifying the ‘practice gap’ in splenic injury management.(13, 18) To use this knowledge to drive practice change, we now need to determine what needs to shift at an individual behavioural and organisational level, and the barriers and facilitators required to achieve these changes.(22) In splenic injury care, the underlying behavioural drivers of different types of surgeons need to be understood. This knowledge is vital to inform the development of interventions that are likely to be acceptable and successful.(23)

In this study, we aimed to identify differences in practice patterns and behavioural drivers in adult and pediatric surgeons related to the management of splenic injury in children aged 0-15 years and young people aged 16-25 years. To do this, we used the COM-B model of behaviour, underpinned by a Theoretical Domains Framework (TDF).(22, 24) The COM-B model integrates a number of behaviour-change theories, grouping domains of Behaviour into three main areas. C: capability, O: opportunity, and M: motivation (Figure 1). The study was approved by the University of New South Wales Human Research Ethics Committee (HC180584).

**Figure 1:**
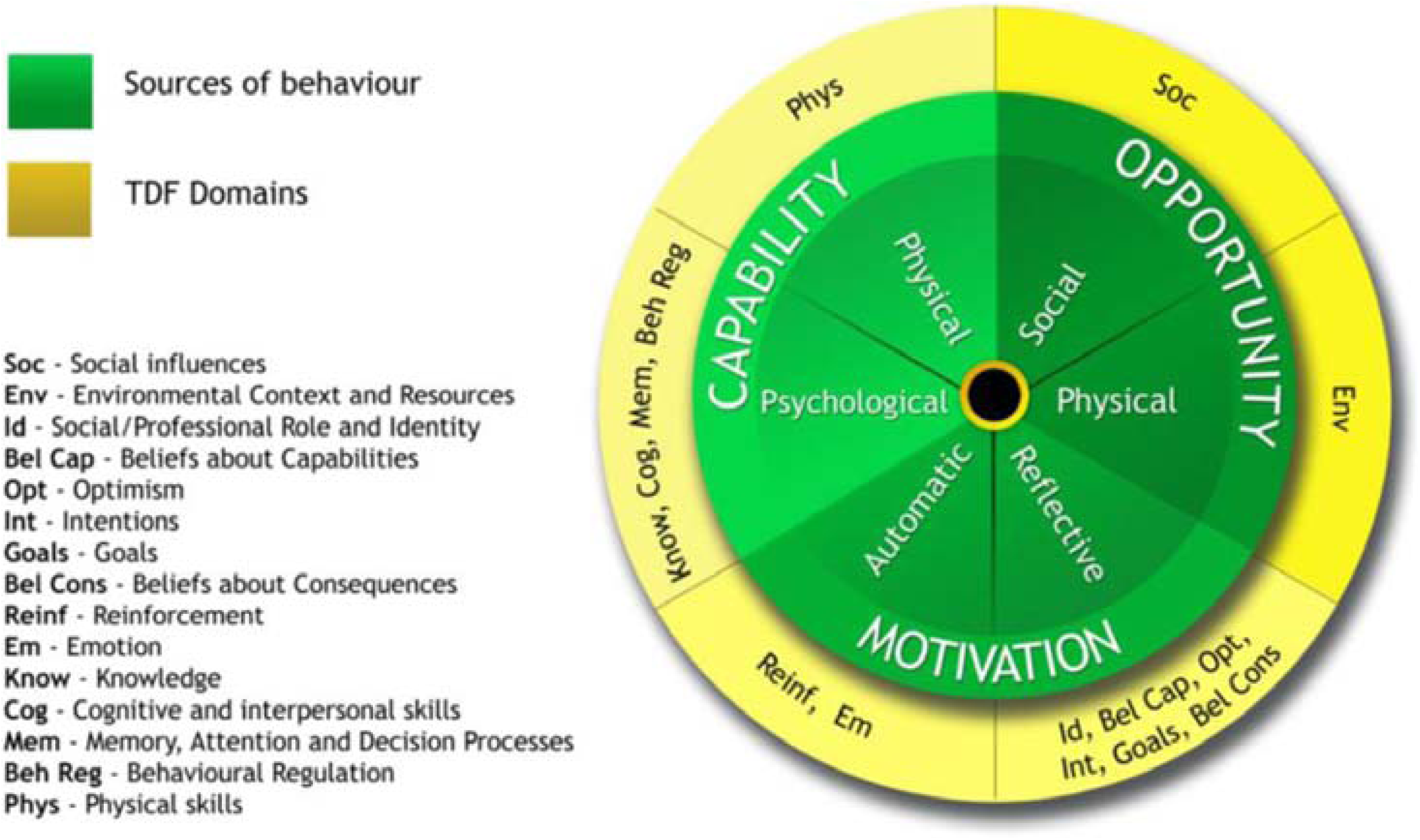
Flow chart illustrating the COM-B basis for designing a questionnaire study. From Atkins (*2017*) A guide to using the Theoretical Domains Framework of behaviour change to investigate implementation problems. With permission under the terms of the Creative Commons Attribution 4.0. (https://creativecommons.org/licenses/by/4.0/) [24]

## METHODS

To achieve our study aim, we used the COM-B model of behaviour and TDF to construct a survey and delivered this to a cross-section of adult general surgeons and pediatric surgeons working in public hospitals throughout NSW, Australia. Data were collected between December 2018 and April 2019.

### Setting

The study was conducted in NSW, Australia. Public hospitals in the state are divided in to (a) major adult trauma centres, of which there are seven, (b) regional/rural trauma centres – comprising ten hospitals throughout regional NSW, and (c) paediatric trauma centres – of which there are three, accepting admissions up to the age of 15 years. One paediatric trauma centre is collocated with a regional trauma centre; the others function independently from the nearby adult facilities. The remaining hospitals are metropolitan or regional/rural local hospitals, neither of which are specifically resourced to deal with major trauma. Depending on location, facility capacity, and injury severity, injured children may be admitted or transferred to paediatric trauma centres, where they are cared for by paediatric surgeons, or remain at adult trauma and non-trauma centres under the care of adult surgeons. There are no specified pre-hospital or interhospital paediatric triage criteria. Previous research in NSW has shown two thirds of children with splenic injury are cared for outside of paediatric trauma hospitals.(13)

### Participants

All adult general surgeons and pediatric surgeons working in NSW public hospitals who were members of the Australia and New Zealand Association of Pediatric Surgeons (ANZAPS) or General Surgeons Australasia (GSA) were invited to participate. Invitations for voluntary participation were emailed via these two organisations. Interested surgeons were required to follow an online link to consent and participate in the survey. Reminder emails were sent at four weeks and ten weeks. Retired surgeons, surgeons working exclusively in a recognised subspecialty, those who never managed trauma, and those in exclusively private practice were excluded. No apriori sample size or composition was set.

### The Survey

The survey was designed for this study and informed by a review of relevant literature,(25–27) the TDF and COM-B model,(22, 24) and discussion with academic and trauma experts. Care was taken to avoid questions that were leading, attempting to assess more than one issue, or negatively phrased. All questions were closed-ended: single option, multiple option, or six-point Likert scale. Responses were anonymous.

Survey questions collected data regarding: **c**apability, including surgeon demographics, qualifications, knowledge, training, and experience; **o**pportunity, including hospital settings and availability of resources and personnel; and **m**otivation, reflected in areas of practice interest and beliefs about optimal care of children and young people with splenic injury. Questions about guidelines covered all three domain categories: familiarity (capability), use (opportunity), and usefulness (motivation) (Table 1).

**Table 1:** Quantitative variables, the survey questions they relate to and their values. (a) Explanatory variables: surgeon characteristics (b) Outcome variables: beliefs and practice patterns. Quantitative variables were created from survey answers. The full survey containing all questions is provided in Appendix 1. PS = pediatric surgeon. AS= adult surgeon. M-AS = metropolitan adult surgeon. R-AS = regional/rural adult surgeon. IQR = interquartile range. PTC = paediatric trauma centre. ATC = adult trauma centre. RTC = regional/rural trauma centre. MLH = metropolitan local hospital. RLH = regional/rural local hospital. FRACS = Fellowship of the Royal Australasian College of Surgeons. EMST = Emergency Management of Severe Trauma. DSTC = Definitive Surgical Trauma Care. APLS = Advanced Paediatric Life Support. OM = operative management. NOM = non-operative management. AE = angioembolisation. *see Supplementary Table 2

| Variable (Data type) | Survey question* | Value | TDF |
| --- | --- | --- | --- |
| **(a) Explanatory Variables: surgeon characteristics** |  |  |  |
| **Surgeon characteristics (survey questions # 2, 3, 6, 16, 17, 18) – Capability, Opportunity** |  |  |  |
| Surgeon type (categorical) – by designation of main public hospital appointment. | #2,3/RACS census (25) | Pediatric (PS)<br>Adult (AS); M-AS, R-AS | Capability: knowledge and skills<br>Motivation: Social/professional role/identity |
| Hospital category (categorical) | # 3 | PTC/ATC/RTC/MLH/RLH | Opportunity: resources, culture, social influences, cues |
| Age - ordinal | # 16 | Years |  |
| Gender - categorical | # 17 | Male/Female |  |
| Australian medical school -categorical | # 18 | Yes/No |  |
| Year of FRACS - ordinal | # 6 | Year (mean, median) | Capability: experience |
| **Surgeon practice (survey questions 4, 5) – Capability, Motivation** |  |  |  |
| Main area of surgical practice - categorical | # 4 | General (AS/PS)/Trauma/Other | Capability: knowledge, experience, behavioural regulation<br>Motivation: Beliefs about capability, optimism |
| Trauma practice interest - categorical | # 4,5 | Yes/No | Capability, Motivation |
| Paediatric practice interest – categorical (AS only) | # 5 | Yes/No | Capability, Motivation |
| **Surgical training and experience (survey questions # 7, 8) - Capability** |  |  |  |
| Trauma related training - categorical | # 8 | EMST/DTSC/APLS/ Trauma fellowship | Capability: experience, skills, knowledge, cognition |
| Training at trauma centre – categorical | # 8 | RTC/ATC/PTC | Capability: experience, skills, knowledge, cognition |
| **(b) Outcome variables: beliefs and practice patterns** |  |  |  |
| **Preference for care of children for elective, emergency and trauma conditions (survey question # 9) - Motivation** |  |  |  |
| Belief about personal skills and capacity – categorical | # 9 | Yes/No | Motivation: beliefs about consequences, optimism, emotions |
| Comfort down to what age – ordinal | # 9 | Years | Motivation - beliefs about consequences, optimism, emotions |
| Comfort with what complexity – ordinal | # 9 | Scale 1-100 | Motivation: beliefs about consequences, optimism, emotions |
| **Experience with splenic injury by age group (survey questions # 10, 11) – Capability, Motivation** |  |  |  |
| Cared for in last 12 months – categorical (0-8, 9-16, 17-25) | # 10 | Yes /No | Capability: experience |
| Preference for place of care – categorical (0-4, 5-8, 9-12, 13-16, 17-19, 20-25) | # 11 | Keep/Transfer | Motivation: intentions, goals, beliefs about consequences. |
| **Guidelines and audit– adult, paediatric (survey question # 12) – Capability, Opportunity, Motivation** |  |  |  |
| Familiarity – categorical | # 12 | Yes/No | Capability: knowledge |
| Use – categorical | # 12 | Yes/No | Opportunity: environmental context, resources |
| Potential/usefulness – categorical | # 12 | Yes/No | Motivation: emotions, goals, beliefs about consequences. |
| Trauma audit involvement - Categorical | # 12 | Yes/No | Opportunity |
| **Available appropriately skilled personnel and resources by age group (survey questions # 13, 14) - Opportunity** |  |  |  |
| Skilled personnel - categorical | # 13 | Yes/No | Opportunity: environmental context, resources, professional support, social influences. |
| Physical resources - categorical | # 14 | Yes/No | Opportunity: resources, |
| **Scenarios 1-10: age 10; age 18 (see Table 2 and supplementary materials for more detail) (survey question # 15) - Behaviour** |  |  |  |
| Preferred management – categorical | # 15 | OM/NOM/AE/Transfer |  |
| Management alignment with pediatric guidelines – categorical | # 15/paediatric guidelines (2) | Yes/No |  |
| Management alignment with adult guidelines – categorical | # 15/adult guidelines (7) | Yes/No |  |

Ten clinical scenarios were developed to collect data on the relationship between practice patterns and surgeon type. In all scenarios, there was a grade IV splenic injury with moderate intraperitoneal blood in 1) a child aged 10 years; and 2) a young adult aged 18 years. Scenarios varied regarding whether there was a contrast blush on CT scan, associated injuries (liver and head), hemodynamic instability, and response to resuscitation. Surgeons were asked to nominate their preferred initial management: operative management, non-operative management, angioembolization, or transfer. These responses were aligned with the most recently published North American guidelines for the management of splenic injury in adults and children. (2, 8) Supplementary Table 1 summarises the scenarios and their alignment with these adult and pediatric guidelines. Key differences between the latter include indications for angioembolization and thresholds for surgery.

An initial draft survey was tested using face-to-face talk-throughs with a small group of representative surgeons (N=4) and experts in survey methodology (n=2). Errors, clinical inaccuracies, omissions, duplications, and inconsistencies were identified and corrected to produce the final survey. The final survey was distributed via Research Electronic Data Capture (REDCap), hosted at the University of NSW, and is provided in Supplementary Table 2.

### Variables

Quantitative variables for analysis were extracted and/or created from survey answers and mapped to TDF in the categories of capability, opportunity, and motivation. They are summarised in Table 1.

Practitioner groups were defined as pediatric or adult surgeons, with adult surgeons sub-grouped into metropolitan and rural. Scaled responses were converted to binary categorical variables: yes or no. Variables constructed from the ten scenarios (Supplementary Table 2: survey questions 15.1 – 15.10) included: preferred management; alignment with the pediatric guidelines (2); and alignment with the adult guidelines,(8) as agreed by two authors (SA and AH). For the purposes of analysing surgeon decision-making in relation to guidelines, the option of ‘transfer’ was considered consistent with both adult and pediatric guidelines for all scenarios.

### Analysis

Open-source R software, version 3.5.2 was used for all analyses.

Descriptive statistics for each variable were generated, with median and interquartile range (IQR) for continuous/ordinal variables and proportions for categorical variables. Completed and partially completed questionnaires were included in the analysis, adjusting the denominator as required.

To identify any response bias, the population of respondents was compared to the wider surgical population as summarised in the Royal Australasian College of Surgeons (RACS) 2018 surgical workforce census.(28) Outcome variables (Table 1) were compared between the practitioner groups. Chi-square or Fisher’s exact tests were used to compare categorical variables. The Wilcoxon rank sum test with continuity correction was used to compare ordinal and continuous variables, as normality was not assumed. A p-value of ≤ 0.05, was accepted as significant.

## RESULTS

Three hundred and three surgeons were invited to participate: 276 (91%) adult surgeons and 27(9%) pediatric surgeons. Ninety-two (30%) responded, with 80 meeting inclusion criteria, including 22 partial responses included for analysis where answers were given. There were 15 pediatric surgeons (19%) and 65 adult general surgeons (81%), with no significant difference in age or year of board certification. The mean age (pediatric surgeon: 50.2 years, adult surgeon: 50.8 years) was younger than the mean age of all surgeons in Australia (57 years).(28)

### Capability

Capability was assessed through training and experience (Table 2: a, b). While 40% of surgeons described trauma as an area of practice interest, only one surgeon from a metropolitan adult trauma centre described it as their main area of practice. Four regional/rural but no metropolitan adult surgeons identified pediatric surgery as an area of interest.

**Table 2:** Comparisons between surgeon types for: (a) area of practice interest; (b) training; (c) experience caring for splenic injury; (d) personnel adequacy; (e) resources adequacy; (f) preference to keep rather than transfer; (g) guideline familiarity, use and usefulness; (h) trauma audit. AS = adult surgeon PS = paediatric surgeon. M = metropolitan. R = regional/rural. EMST = Emergency Management of Severe Trauma. APLS = Advanced Paediatric Life Support DSTC = Definitive Surgical Trauma Care. TF = trauma fellowship. SET= surgical education and training. TF = trauma fellowship. AG = adult guideline. PG = paediatric guideline.

| Surgeon type<br>N=76 | AS<br>N=61(%) | PS<br>N=15(%) | p | M-AS<br>N=27(%) | R-AS<br>N=34(%) | p |
| --- | --- | --- | --- | --- | --- | --- |
| (a) Practice interest |  |  |  |  |  |  |
| General Surgery | 30(49) | 11(73) | 0.14 | 6(22) | 24(71) | <0.001 |
| Trauma | 25(41) | 5 (33) | 0.77 | 13(48) | 12(35) | 0.43 |
| (b) training |  |  |  |  |  |  |
| EMST | 55(92) | 12(80) | 0.19 | 23(88) | 32(94) | 0.64 |
| APLS | 5(8) | 12(80) | **<0.001** | 1(4) | 4(12) | 0.38 |
| DSTC | 36(60) | 8(53) | 0.77 | 15(58) | 21(62) | 0.79 |
| TF | 1(2) | 1(7) | 0.37 | 1(4) | 0(0) | 0.44 |
| SET at ATC | 44(73) | 9(60) | 0.35 | 23(88) | 21(62) | 0.04 |
| SET at PTC | 9(15) | 15(100) | **<0.001** | 2(8) | 7(21) | 0.27 |
| SET at RTC | 30(50) | 7(47) | 1.00 | 12(46) | 18(53) | 0.79 |
| (c) experience |  |  |  |  |  |  |
| Age 0-8 | 9(16) | 12(80) | **<0.001** | 1(4) | 7(22) | 0.12 |
| Age 9-16 | 25(45) | 12(80) | **<0.05** | 7(30) | 18(56) | 0.10 |
| Age 17-25 | 37(66) | 1(8) | **<0.001** | 17(74) | 20(62) | 0.39 |
| Surgeon<br>Type N=58 | AS<br>N=45(%) | PS<br>N=13(%) | p | M-AS<br>N=17(%) | R-AS<br>N=28(%) | p |
| (d) adequate personnel |  |  |  |  |  |  |
| 0-4 | 13(29) | 13(100) | **<0.001** | 6(35) | 7(25) | 0.51 |
| 5-8 | 17(38) | 13(100) | **<0.001** | 6(35) | 11(39) | 1.00 |
| 9-12 | 25(56) | 13(100) | **<0.01** | 9(53) | 16(57) | 1.00 |
| 13-16 | 38(84) | 13(100) | 0.32 | 15(88) | 23(82) | 1.00 |
| 17-19 | 43(96) | 9(69) | **<0.01** | 17(100) | 26(93) | 1.00 |
| 20-25 | 44(98) | 5(38) | **<0.001** | 17(100) | 27(96) | 1.00 |
| (e) adequate resources |  |  |  |  |  |  |
| 0-4 | 12(27) | 13(100) | **<0.001** | 8(47) | 4(14) | **<0.05** |
| 5-8 | 15(33) | 13(100) | **<0.001** | 8(47) | 7(25) | 0.19 |
| 9-12 | 20(44) | 13(100) | **<0.001** | 11(65) | 9(32) | 0.07 |
| 13-16 | 34(76) | 13(100) | 0.11 | 15(88) | 19(68) | 0.16 |
| 17-19 | 40(89) | 10(77) | 0.36 | 16(94) | 24(86) | 0.63 |
| 20-25 | 41(91) | 4(31) | **<0.001** | 16(94) | 25(89) | 1.00 |
| Surgeon type N=71 | AS<br>N=56(%) | PS<br>N=15(%) | p | M-AS<br>N=23(%) | R-AS<br>N=33(%) | p |
| (f) preference to keep rather than transfer |  |  |  |  |  |  |
| Age 0-4 | 11(20) | 15(100) | **<0.001** | 4(17) | 7(21) | 1.00 |
| Age 5-8 | 20(36) | 15(100) | **<0.001** | 6(26) | 14(42) | 0.26 |
| Age 9-12 | 29(52) | 15(100) | **<0.001** | 8(35) | 21(64) | 0.06 |
| Age 13-16 | 45(80) | 15(100) | 0.10 | 16(70) | 29(88) | 0.16 |
| Age 17-19 | 52(93) | 7(47) | **<0.001** | 20(87) | 32(97) | 0.29 |
| Age 20-25 | 53(95) | 6(40) | **<0.001** | 21(87) | 32(97) | 0.56 |
| Surgeon type (N=58) | AS<br>N=45(%) | PS<br>N=13(%) | p | M-AS<br>N=17(%) | R-AS<br>N=28(%) | p |
| (g) guideline familiarity, use and potential, and (h) trauma audit involvement |  |  |  |  |  |  |
| AG Familiarity | 32(71) | 4(31) | **<0.05** | 12(71) | 20(71) | 1.00 |
| AG Use | 28(62) | 3(23) | **<0.05** | 10(59) | 18(64) | 1.00 |
| AG Potential | 42(93) | 12(92) | 1.00 | 17(100) | 25(89) | 0.28 |
| PG Familiarity | 16(36) | 13(100) | **<0.001** | 5(29) | 11(39) | 0.50 |
| PG Use | 14(31) | 11(85) | **<0.001** | 2(12) | 12(43) | **0.05** |
| PG Potential | 41(91) | 13(100) | 0.56 | 16(94) | 25(89) | 1.00 |
| Adult audit | 28(62) | 1(8) | **<0.001** | 11(65) | 17(61) | 1.00 |
| Paediatric audit | 10(22) | 7(54) | **<0.05** | 1(6) | 9(32) | 0.06 |

Just under three-quarters of surgeons (n= 53, 70%) undertook at least part of their surgical education and training (SET) at a metropolitan adult trauma centre, with no significant difference between adult and pediatric surgeons. Fewer adult surgeons at regional/rural hospitals than at metropolitan hospitals had such a placement (p<0.05). All pediatric surgeons undertook at least part of their SET at a pediatric trauma centre, but less than one-fifth (n=9, 15%) of adult surgeons had this experience (p<0.001).

Emergency Management of Severe Trauma (EMST, equivalent to Advanced Trauma Life Support), was undertaken by almost 90% of respondents, and Definitive Surgical Trauma Care (DSTC) was completed by just over half (59%), with no significant difference between pediatric and adult surgeons. The only specifically pediatric course containing trauma training was Advanced Pediatric Life Support (APLS, equivalent to Pediatric Advanced Life Support), which was completed by significantly more pediatric than adult surgeons (p<0.001).

The majority (80%) of pediatric surgeons had cared for a case of pediatric splenic injury in the last 12 months, largely confined to patients under the age of 17. For adult surgeons, this increased with the age of the child (18% age 0-8 up to 46% age 9-16). Two-thirds of adult surgeons had cared for a young adult with splenic injury in the last 12 months (Table 2: c)

### Opportunity

Hospital location and availability of resources and personnel reflected the opportunity to care for splenic injury. Over half of the respondents were from regional/rural hospitals (55%), compared with 30% reported in the RACS 2018 census.(28) Twelve pediatric surgeons (80%) worked in pediatric trauma centres. Of the adult surgeons, 19(29%) worked in metropolitan adult trauma centres, 24 (37%) in regional/rural trauma centres, 10 (15%) in metropolitan local hospitals, and 12 (19%) in regional/rural local hospitals (Table 2: a).

Significantly more pediatric than adult surgeons believed that personnel and resources in their practice setting were adequate to allow the opportunity for the care of children up to the teenage years, but there was no significant difference for ages 13-16. In young adults, significantly more adult that pediatric surgeons felt personnel were adequate over the age of 16, and resources adequate over the age of 19 (Table 2: d, e)

### Motivation

All pediatric surgeons believed they had the skills and capacity to care for children in elective, emergency, and trauma settings at all ages. Three-quarters of adult surgeons were comfortable with elective care of children, rising to around 90% in an emergency or trauma situation. There were significant caveats on the lower age limit and degree of complexity. Rural surgeons were significantly more comfortable down to a lower age than their metropolitan counterparts in all three areas (Table 3).

**Table 3:** Motivation: Comfort with the elective, emergency and trauma care of children. Comparing (a) adult and paediatric surgeons and (b) regional/rural and metropolitan adult surgeons. AS = adult surgeon. PS = paediatric surgeon. M = metropolitan. R = regional/rural. Complexity reported by surgeons on a sliding scale from 0-100.

**(a) Adult and paediatric surgeons**
| Surgeon type | Elective |  |  | Emergency |  |  | Trauma |  |  |
| --- | --- | --- | --- | --- | --- | --- | --- | --- | --- |
|  | AS<br>N=58(%) | PS<br>N=15(%) | p | AS<br>N=58(%) | PS<br>N=15(%) | p | AS<br>N=58(%) | PS<br>N=15(%) | p |
| Skills and capacity to care for children | 44(76) | 15(100) | 0.06 | 53(91) | 15(100) | 0.58 | 51(88) | 15(100) | 0.33 |
| If yes – down to what age (years) |  |  |  |  |  |  |  |  |  |
| Median | 5.0 | 0 | **<0.001** | 3.0 | 0 | **<0.001** | 4.0 | 0 | **<0.001** |
| IQR | 2.5-5.5 | - |  | 2-5 | 0 |  | 2-5 | 0 |  |
| Complexity (scale 1-100) |  |  |  |  |  |  |  |  |  |
| Median | 32.5 | 100.0 | **<0.001** | 38.2 | 100.0 | **<0.001** | 40.0 | 99.0 | **<0.001** |
| IQR | 18-51 | 98-100 |  | 28-51 | 94-100 |  | 26-50 | 85-100 |  |

**(b) Regional/rural and metropolitan adult surgeons**
| Surgeon type | M-AS<br>N=25(%) | R-AS<br>N=33(%) | p<br>OR | M-AS<br>N=25(%) | R-AS<br>N=33(%) | p | M-AS<br>N=25(%) | R-AS<br>N=33(%) | p |
| --- | --- | --- | --- | --- | --- | --- | --- | --- | --- |
| Skills and capacity to care for children | 18(72) | 26 (79) | 0.76 | 20 (80) | 33(100) | <0.05 | 19 (76) | 32 (97) | <0.05 |
| If yes – down to what age (years) |  |  |  |  |  |  |  |  |  |
| Median | 5.5 | 3.0 | **<0.01** | 5.0 | 3.0 | **<0.001** | 6.0 | 3.0 | **<0.001** |
| IQR | 4.2-10 | 2-5 |  | 4-10 | 2-4 |  | 4-10 | 2-5 |  |
| Complexity (scale 0-100) |  |  |  |  |  |  |  |  |  |
| Median | 34.0 | 31.5 | 0.76 | 42.5 | 42.0 | 0.96 | 40.0 | 40.0 | 0.93 NS |
| IQR | 14-51 | 23-50 |  | 28-51 | 28-50 |  | 26-51 | 26-50 |  |

Adult surgeon preference for transfer of children with splenic injury was strong for the very young patient, although not universal (Table 2: f). This decreased with increasing age of the child. Eighty percent of adult surgeons preferred to transfer rather than keep a child <four years and 64% between five and eight years. This decreased to 50% for children > nine years, and 20% for children aged > 13 years. Comparing regional/rural, and metropolitan adult surgeons, transfer preference was lower among the former for children of all ages, although this did not reach significance: two-thirds (65%) of adult surgeons in metropolitan areas preferred transfer of children 9-12 compared to just over a third (36%) in regional/rural areas (p=0.06).

### Guidelines

Beliefs regarding guidelines and audits were reflective of capability (guideline familiarity), opportunity (guidelines in use), and motivation (beliefs about guideline usefulness) (Table 2: g). Adult surgeons were more likely to be familiar with and report the use of adult guidelines (p<0.05), but less likely to be familiar with and report the use of pediatric guidelines (p<0.001). Regardless of familiarity and reported use, belief among all surgeons was high (>90%) that both adult and pediatric guidelines have the potential to improve care. Involvement in trauma audits was not universal, at just over 50% of respondents. It was particularly low for adult surgeons regarding audit of pediatric trauma (p<0.05) and pediatric surgeons regarding audit of adult trauma (p<0.001).

### Management patterns

Scenario responses are presented in detail in Supplementary Table 3: a – f, with Table 4 tracking them to pediatric and adult guidelines.(2, 8) There were differences in management preferences between adult and pediatric surgeons, but no significant difference was observed between regional/rural and metropolitan adult surgeons for any scenario. Differences in preferred initial management by surgeon type were observed depending upon: the presence of contrast blush on CT scan, the extent of hemodynamic instability, and the presence of associated injuries – in particular, a head injury.

**Table 4:** Tracking scenario responses to splenic injury management guidelines by surgeon type. (a) Paediatric (ATOMAC) guidelines (b) Adult (WTA) guidelines. All scenarios included a grade IV splenic injury with moderate intraperitoneal blood. Pediatric guidelines: ATOMAC = the North American Paediatric Trauma Consortium [2]. Adult guidelines: WTA = Western Trauma Association [8]. AS = adult surgeon. PS = paediatric surgeon. CB = contrast blush. TBI = traumatic brain injury. PS = paediatric surgeon. AS = adult surgeon. *see Supplementary Tables 1 and 2

| (a) Tracking to pediatric guidelines [2] |  |  |  |  |  |  |  |  |  |  |  |  |  |  |
| --- | --- | --- | --- | --- | --- | --- | --- | --- | --- | --- | --- | --- | --- | --- |
| No contrast blush |  |  | Associated issues |  |  |  | PS n (%) |  |  | AS n (%) |  |  | p – AS vs PS |  |
| Scenario* | N PS/AS | recommendation | CB | unstable | TBI | Liver | Age 10 | Age 18 | p | Age 10 | Age 18 | p | Age 10 | Age 18 |
| 1 | 15/53 | NOM/TRANS | - | - | - | - | 15(100) | 15(100) | 1.00 | 51(96) | 50(94) | 1.00 | 1.00 | 1.00 |
| 4 | 14/49 | NOM/TRANS | - | - | - | + | 14(100) | 14(100) | 1.00 | 49(100) | 46(94) | 0.24 | 1.00 | 1.00 |
| 7 | 14/47 | NOM/TRANS | - | - | + | - | 14(100) | 14(100) | 1.00 | 46(98) | 44(94) | 0.62 | 1.00 | 1.00 |
| 8 | 14/47 | NOM/TRANS | - | + | + | - | 13(93) | 14(100) | 1.00 | 46(98) | 42(89) | 0.20 | 0.40 | 0.58 |
| Contrast blush |  |  | Associated issues |  |  |  | PS n (%) |  |  | AS n (%) |  |  | p – AS vs PS |  |
| Scenario* | N PS/AS | recommendation | CB | unstable | TBI | Liver | Age 10 | Age 18 | p | Age 10 | Age 18 | p | Age 10 | Age 18 |
| 2 | 15/52 | NOM/TRANS | + | - | - | - | 12(80) | 11(73) | 1.00 | 33(63) | 16(31) | **<0.01** | 0.35 | **<0.01** |
| 3 | 14/49 | NOM/TRANS | + | + | - | - | 11(79) | 10(71) | 0.67 | 30(61) | 12(25) | **<0.001** | 0.34 | **<0.01** |
| 5 | 14/48 | NOM/TRANS | + | - | - | + | 10(71) | 8(57) | 0.69 | 36(75) | 15(31) | **<0.001** | 1.00 | 0.12 |
| 6 | 14/48 | NOM/AE/TRANS | + | ++ | - | + | 13(93) | 12(86) | 1.00 | 31(65) | 22(46) | 0.10 | **<0.05** | **<0.05** |
| 9 | 14/47 | NOM/AE/TRANS | + | ++ | - | - | 13(93) | 12(86) | 1.00 | 31(66) | 25(53) | 0.29 | 0.08 | **<0.05** |
| No contrast blush, ongoing instability |  |  | Associated issues |  |  |  | PS n (%) |  |  | AS n (%) |  |  | p – AS vs PS |  |
| Scenario* | N PS/AS | recommendation | CB | unstable | TBI | Liver | Age 10 | Age 18 | p | Age 10 | Age 18 | p | Age 10 | Age 18 |
| 10 | 14/46 | NOM/AE/OM/TRANS | - | +++ | + | + | 15(100) | 46(100) | 1.00 | 46(100) | 46(100) | 1.00 | 1.00 | 1.00 |
| (b) Tracking to adult guidelines [8] |  |  |  |  |  |  |  |  |  |  |  |  |  |  |
| No contrast blush |  |  | Associated issues |  |  |  | PS n (%) |  |  | AS n (%) |  |  | p – AS vs PS |  |
| Scenario* | N PS/AS | recommendation | CB | unstable | TBI | Liver | Age 10 | Age 18 | p | Age 10 | Age 18 | p | Age 10 | Age 18 |
| 1 | 15/53 | NOM/AE/TRANS | - | - | - | - | 15(100) | 15(100) | 1.00 | 52(98) | 52(98) | 1.00 | 1.00 | 1.00 |
| 4 | 14/49 | NOM/AE/TRANS | - | - | - | + | 14(100) | 14(100) | 1.00 | 49(100) | 49(100) | 1.00 | 1.00 | 1.00 |
| 7 | 14/47 | NOM/AE/TRANS | - | - | + | - | 14(100) | 14(100) | 1.00 | 47(100) | 46(98) | 0.61 | 1.00 | 1.00 |
| 8 | 14/47 | AE/TRANS | - | + | + | - | 3(21) | 4(29) | 1.00 | 39(83) | 25(53) | <0.01 | <0.001 | 0.13 |
| Contrast blush |  |  | Associated issues |  |  |  | PS n (%) |  |  | AS n (%) |  |  | p – AS vs PS |  |
| Scenario* | N PS/AS | recommendation | CB | unstable | TBI | Liver | Age 10 | Age 18 | p | Age 10 | Age 18 | p | Age 10 | Age 18 |
| 2 | 15/52 | AE/TRANS | + | - | - | - | 3(20) | 8(53) | 0.13 | 43(88) | 38(78) | 0.34 | **<0.001** | 0.2 |
| 3 | 14/49 | AE/TRANS | + | + | - | - | 4(29) | 8(57) | 0.25 | 39(80) | 33(67) | 0.25 | **<0.001** | 0.53 |
| 5 | 14/48 | AE/TRANS | + | - | - | + | 4(29) | 9(64) | 0.12 | 45(94) | 41(85) | 0.31 | **<0.001** | 0.12 |
| 6 | 14/48 | AE/OM/TRANS | + | ++ | - | + | 12(86) | 12(86) | 1.00 | 48(100) | 47(98) | 1.00 | **<0.05** | 0.12 |
| 9 | 14/47 | AE/OM/TRANS | + | ++ | - | - | 12(86) | 12(86) | 1.00 | 47(97) | 46(97) | 1.00 | 0.14 | 0.13 |
| No contrast blush, ongoing instability |  |  | Associated issues |  |  |  | PS n (%) |  |  | AS n (%) |  |  | p – AS vs PS |  |
| Scenario* | N PS/AS | recommendation | CB | unstable | TBI | Liver | Age 10 | Age 18 | p | Age 10 | Age 18 | p | Age 10 | Age 18 |
| 10 | 14/46 | OM/TRANS | - | +++ | + | + | 5(36) | 8(57) | 0.45 | 42(91) | 39(85) | 0.52 | **<0.001** | 0.06 |

In a stable patient with an absent contrast blush (Scenarios 1, 4, 7, and 8; Supplementary Table 3: a, b), operative management was rare (less than 5%) and use of angioembolization, low (less than 6%), with most surgeons choosing non-operative management or transfer. Correlation with both adult and pediatric guidelines was high for all surgeons, except for scenario 8, where adult guidelines recommend embolization. Transfer was common in the presence of a head injury from centres with no neurosurgery.

When a contrast blush was present (Supplementary Table 3: c, d), most pediatric surgeons did not routinely intervene in the stable patient (Scenarios 2,3 and 5). However, up to one third considered angioembolization, more aligned with adult rather than pediatric guidelines. They increasingly intervened in the more unstable the patient (Scenarios 8, 10), with angioembolization frequently chosen ahead of operation.

In general, adult surgeons moved more readily than pediatric surgeons from a preference for angioembolization in the presence of a contrast blush, to operation in the presence of any instability. This is consistent with adult rather than pediatric guidelines. The exception was a higher propensity among adult surgeons to transfer a child, rather than intervening, particularly in the presence of a head injury. However, there were some situations where adult surgeons intervened less than the adult guidelines advise, including persisting with non-operative management in the patient with no contrast blush but requiring one lot of resuscitation (Scenario 8), and choosing operation ahead of angioembolization in the relatively stable patient (Scenarios 2 and 3). Adult surgeons appeared to adjust management choice based on patient age, with higher intervention rates in young adults compared to children, particularly higher use of angioembolization in the presence of a contrast blush.

Depending on the scenario, 46-81% of adult surgeons preferred to transfer the 10-year-old. Transfer was chosen more often in the context of an extradural haemorrhage (Scenario 7: 37/47, 79%; Scenario 8: 38/47, 81%). This was countered by a lower inclination to transfer the very unstable child (scenario 10: 21/46, 46%).

## DISCUSSION

This survey has identified differences between adult and surgeons in their management of splenic injury and is the first study to attempt to systematically examine the behavioural drivers responsible for these observed differences.

Our finding that simpler cases are often managed non-operatively by all surgeons is consistent with the findings of other studies.(26, 29) However, divergence in preference between adult and paediatric surgeons was evident with increasing scenario complexity due to contrast blush, associated injuries, and hemodynamic instability. Research internationally has also shown that adult surgeons, concerned about non-operative management failure, are more likely to intervene, either with operation or embolisation, in both in children (16, 27) and young adults.(17, 19, 30, 31)

Importantly, this study has identified drivers of these variations in management decisions beyond surgeon and hospital type, that can be mapped to known domains of behaviour (Figure 1).(22, 24) Differences between adult and pediatric surgeons regarding training and experience in pediatric surgery and trauma management appear to drive ‘capability’ aspects of behaviour. Adult surgeons have less exposure to pediatric trauma during SET and are less likely to undergo APLS training.

For ‘opportunity’ in terms of personnel and resources, more pediatric than adult surgeons believed the resources and personnel in their workplace were adequate to care for very young children. Only one-third of adult surgeons felt the same, with their optimism increasing with the age of the child. This suggests that the opportunity to deliver evidence-based care is enhanced by personnel with expertise, in an environment that focuses on the specific physical and psychological needs of children. Resources and personnel may include paediatric colleagues, nursing, intensive care, operating theatres, and access to interventional radiology.

Management guidelines in use in different settings may also contribute to the opportunity to provide evidence-based care: adult surgeons are familiar with and use adult guidelines, while pediatric surgeons are familiar with and use pediatric guidelines. This correlates with the more frequent choice of angioembolisation by adult surgeons in the presence of a contrast blush. In adult guidelines, angioembolisation is seen and an adjunct to non-operative management, generally reserved for the stable patient, prompted by grade, haemoperitoneum, contrast blush on CT-scan and/or evidence of ongoing bleeding.(8) In contrast, paediatric guidelines do not recommend any intervention in the stable child, regardless of grade or contrast blush. Instead, angioembolisation is not mandated but is considered alongside operative management for recurrent haemodynamic instability and falling haemoglobin.(2) Studies have shown a higher use of angioembolisation at adult trauma centres than paediatric trauma centres in the management of paediatric and adolescent splenic injury.(15–17) It is unclear if this is due to availability or choice, and whether its use increases splenic salvage in children.(15, 16, 31) This requires further exploration.

‘Motivation’ as a driver of behaviour is reflected in the surgeons’ reported comfort with the care of children in the elective, emergency and trauma situations. While more than three-quarters of adult surgeons expressed such comfort, they placed caveats on age and complexity, with rural surgeons generally comfortable down to a younger age or patient. The latter is consistent with a rural surgeon’s more generalist practice compared to their metropolitan counterparts, and is also driven by the psychosocial and financial needs of families and the practicalities of children otherwise having to travel vast distances to receive care.

The insights into surgeon behavioural drivers of splenic injury management decisions for children and young patients provide potential avenues for addressing previously described disparities in care.(13, 18) Our results indicate that increasing access for adult surgeons to pediatric trauma training and experience may be important. Once in consultant clinical practice, accessing quality evidence, keeping knowledge and skills up to date, and clinical exposure to case numbers can influence management choices.(32) This has implications for surgical training programs as well as the design and content of trauma courses and ongoing professional development throughout a surgeon’s career.

More broadly, our results demonstrate the interaction between clinical guidelines and other behavioural drivers of management decisions. It is encouraging to see the optimism surgeons expressed for the value of guidelines in improving clinical practice. Guideline use has been correlated with the adoption of practice change in other settings.(33, 34) In developing countermeasures to reduce disparity in the care of children and young people with splenic injury, there is a need to ensure that the guidelines being used are appropriate for the setting. This will help ensure continuity of best practice care for a child or young adult of any age at any hospital type, not just in adult or pediatric trauma centres. The work of Giusti (2016) supports this, suggesting that parachuting in guidelines from adult settings to pediatric settings and vice versa is unlikely to be successful, whereas working with surgeons in each of the different settings to extend or adapt guidelines to their own contexts may be more worthwhile.(35)

Confidence in the adequacy of existing resources is also likely to be affected by the physical environment, (35) the facility type and its geographic location,(36) organisational culture, (35) and leadership,(37) as well as the opportunity to connect with professional support.(32) Harnessing local resources and personnel is important in any intervention developed to address care disparities. This needs to be explored in detail in future research.

### Limitations

The questionnaire was not piloted because of the relatively small numbers available to survey, particularly pediatric surgeons. Instead, the survey was refined using ‘talk-through’, as described in the methods section. Despite the wide invitation, the response rate was low at only 30%, with 26% available for analysis after exclusion. However, this was in line with the response rates in similar studies of 23-38%.(27, 29) A low response rate means the study was subject to non-response bias. This was addressed by analysing surgeons in groups as pediatric or adult surgeons, as well as comparing adult regional/rural and metropolitan surgeons. Statistical methods appropriate for small numbers and non-normally distributed data were used. Surgeon characteristics such as training and experience tended to correlate with the surgeon type (pediatric or adult), allowing some conclusions about their relevance to be drawn. Management choices were also consistent with what has been observed at a population level in our previous research.(13, 18) Surgeon responses may also be affected by acquiescence and conformity bias. This may explain the strong correlation of paediatric and adult surgeon management choices with paediatric and adult guidelines respectively. These biases were minimised by anonymity, scaled responses where appropriate, avoiding leading questions, and the variety of clinical scenarios presented.

The survey was long which added to the richness of the data, but may have contributed to higher levels of partial completion. Further targeted research, qualitatively exploring more deeply the factors leading surgeons to pursue certain management strategies would be useful.

## Conclusion

This study has confirmed variations in the management of splenic injury in children and young people between adult and pediatric surgeons. As surgeons are key stakeholders in the delivery of trauma care, understanding the barriers and facilitators driving these variations is a critical prelude to developing and sustainably implementing evidence-based changes to practice.

The results indicate that contextually relevant guidelines for managing splenic injury in children and young people across any setting may be needed to reduce disparities in care. These should be underpinned by interventions designed to further address the drivers of surgeon behaviour to optimise uptake. This includes appropriate training and experience (enhanced capability), an adequate workplace environment, including required resources and personnel at a service level (optimised opportunity), and engaging surgeons in their use (increased motivation).

Finally, it is important to note that splenic injury management is an indicator of trauma care quality, and implementing changes that improve it has the potential to translate to other improvements in trauma care.

## Data Availability

All data produced in the present study are available upon reasonable request to the authors

