## Supplemental Tables 1-3 for "Management of splenic injury in children and young adults: a survey of surgeons investigating factors responsible for variations in care using the Theoretical Domains Framework"

| Supplementary Table 1: Scenarios grouped and tracked to paediatric and adult splenic injury management guidelines recommendations  Paediatric guidelines = ATOMAC [Notrica, 2017 #2183]. Adult guidelines = WTA [Rowell, 2017 #2162]. All scenarios are about (a) a 10-year-old and (b) an 18-year-old with a grade IV splenic injury. They are in three groups: (1) No contrast blush on CT-scan; (2) Contrast blush from the spleen on CT-scan; and (3) Increasing instability, no contrast bush on CT-scan. Management options consistent with guideline recommendations are shaded. Pediatric guideline: ATOMAC = the North American Paediatric Trauma Consortium (Arizona-Texas-Oklahoma-Memphis-Arkansas). Adult guideline: WTA = Western Trauma Association. NOM=non-operative management. OM=operative management. AE=angioembolisation. TRANS=transfer. | | | |
| --- | --- | --- | --- |
| **Number** | **Description** | **pediatric** | **adult** |
| Group 1: No contrast blush on CT-scan | | | |
| # 15.1 | Grade IV isolated splenic injury.  Haemodynamically STABLE on admission.  Moderate intraperitoneal blood, with NO CONTRAST EXTRAVASATION (‘blush’) on abdominal CT-scan. | NOM | NOM |
|  |  | OM | OM |
|  |  | AE | AE |
|  |  | TRANS | TRANS |
| # 15.4 | Grade IV splenic injury WITH GRADE II LIVER LACERATION.  Haemodynamically STABLE on admission.  Moderate intraperitoneal blood, with NO CONTRAST EXTRAVASATION. (‘blush’) on abdominal CT-scan. | NOM | NOM |
|  |  | OM | OM |
|  |  | AE | AE |
|  |  | TRANS | TRANS |
| # 15.7 | Grade IV splenic injury INTUBATED AND VENTILATED.  SMALL EXTRADURAL HAEMATOMA on head CT-scan.  Haemodynamically STABLE on admission. Moderate intraperitoneal blood, with NO CONTRAST EXTRAVASATION (‘blush’) on abdominal CT-scan. | NOM | NOM |
|  |  | OM | OM |
|  |  | AE | AE |
|  |  | TRANS | TRANS |
| # 15.8 | Grade IV splenic injury. INTUBATED AND VENTILATED.  SMALL EXTRADURAL HAEMATOMA on head CT-scan.  Surgeon suspects intra-abdominal bleeding on admission.  Sustained response to 10mls/kg normal saline and transfusion of 10mls/kg packed red blood cells (10-year-old) saline bolus and transfusion of 2 units packed cells (18-year-old).  Moderate intraperitoneal blood, with NO CONTRAST EXTRAVASATION (‘blush’) on abdominal CT-scan. | NOM | NOM |
|  |  | OM | OM |
|  |  | AE | AE |
|  |  | TRANS | TRANS |
| Group 2: Contrast blush from the spleen on CT-scan | | | |
| # 15.2 | Grade IV isolated splenic injury.  Haemodynamically STABLE on admission.  Moderate intraperitoneal blood, with CONTRAST EXTRAVASATION (‘blush’) PRESENT on abdominal CT-scan (from the spleen). | NOM | NOM |
|  |  | OM | OM |
|  |  | AE | AE |
|  |  | TRANS | TRANS |
| # 15.3 | Grade IV isolated splenic injury.  Surgeon suspects intra-abdominal bleeding on admission.  Sustained response to10mls/kg of normal saline and transfusion of 10mls/kg packed red blood cells (10-year-old) saline bolus and transfusion of 2 units packed red blood cells (18-year-old).  Moderate intraperitoneal blood, with CONTRAST EXTRAVASATION (‘blush’) PRESENT on abdominal CT-scan (from the spleen). | NOM | NOM |
|  |  | OM | OM |
|  |  | AE | AE |
|  |  | TRANS | TRANS |
| # 15.5 | Grade IV splenic injury WITH GRADE II LIVER LACERATION.  Haemodynamically STABLE on admission.  Moderate intraperitoneal blood, with CONTRAST EXTRAVASATION (‘blush’) PRESENT on abdominal CT-scan (from the spleen). | NOM | NOM |
|  |  | OM | OM |
|  |  | AE | AE |
|  |  | TRANS | TRANS |
| Continued next page | | | |

| # 15.6 | Grade IV splenic injury WITH GRADE II LIVER LACERATION.  Surgeon suspects intra-abdominal bleeding on admission.  Initial response to 10mls/kg normal saline and transfusion of 10mls/kg packed red blood cells (10-year-old) saline bolus plus transfusion of 2 units packed red blood cells (18-year-old). Develops recurrent haemodynamic instability at 4 hours which responds to further appropriate blood product administration. Moderate intraperitoneal blood, with CONTRAST EXTRAVASATION (‘blush’) PRESENT on abdominal CT-scan (from the spleen). | NOM | NOM |
| --- | --- | --- | --- |
|  |  | OM | OM |
|  |  | AE | AE |
|  |  | TRANS | TRANS |
| # 15.9 | Grade IV splenic injury. INTUBATED AND VENTILATED.  SMALL EXTRADURAL HAEMATOMA on head CT-scan.  Surgeon suspects intra-abdominal bleeding on admission.  Responds initially to 10mls/kg normal saline and transfusion of 10mls/kg packed red blood cells (10-year-old) saline bolus and transfusion of 2 units packed cells (18-year-old). Develops recurrent haemodynamic instability at 4 hours which responds to further appropriate blood product administration. Moderate intraperitoneal blood, and CONTRAST EXTRAVASATION (‘blush’) PRESENT on abdominal CT-scan (from the spleen). | NOM | NOM |
|  |  | OM | OM |
|  |  | AE | AE |
|  |  | TRANS | TRANS |
| Group 3: Increasing instability, no contrast bush on CT-scan | | | |
| # 15.10 | Grade IV splenic injury WITH GRADE II LIVER LACERATION.  INTUBATED AND VENTILATED.  SMALL EXTRADURAL HAEMATOMA on head CT-scan.  Surgeon suspects intra-abdominal bleeding on admission.  Responds initially to 10mls/kg saline and transfusion of 10mls/kg packed red blood cells (10-year-old) saline bolus and transfusion of 2 units packed red blood cells (18-year-old). Develops recurrent haemodynamic instability at 4 hours, which transiently responds to further appropriate blood product administration. Haemodynamic instability recurs over the following 2 hours, which is, again, appropriately resuscitated. Moderate intraperitoneal blood, and NO CONTRAST EXTRAVASATION (‘blush’) on abdominal CT-scan. | NOM | NOM |
|  |  | OM | OM |
|  |  | AE | AE |
|  |  | TRANS | TRANS |

| Supplementary Table 2: Survey  Questions that appeared as a result of branching logic are in italics. Questions asked only of adult surgeons are marked with the suffix -AS. Questions asked only of paediatric surgeons are marked with the suffix -PS. OM = operative management. NOM = non-operative management. AE = angioembolisation | | |
| --- | --- | --- |
|  | Question | Option |
| 1 | Participant information - Please take the time to read the attached information sheet about the study, then indicate in the question below whether or not you wish to take part in the survey. | 1, I have read the information and I agree to participate  2, I wish to decline to participate |
| 2 | Do you undertake some or all of your practice in a PUBLIC HOSPITAL in NSW? | 1, Yes, as an ADULT GENERAL SURGEON (with or without a subspecialty)  2, Yes, as a PAEDIATRIC GENERAL SURGEON (with or without a subspecialty)  3, No, I am RETIRED  4, No, I am ONLY IN PRIVATE PRACTICE |
| 3 | Click on the hospital group that best describes the MAIN PUBLIC HOSPITAL that you work at. | 1, ADULT MAJOR TRAUMA SERVICE (John Hunter Hospital, St George Hospital, Royal Prince Alfred Hospital, Royal North Shore Hospital, St Vincent's Hospital, Liverpool Hospital, Westmead Adult Hospital)  2, REGIONAL TRAUMA SERVICE (Coffs Harbour Base Hospital, Gosford Hospital, Lismore Base Hospital, Nepean Hospital, Orange Health Service, Port Macquarie Base Hospital, Tamworth Rural Referral Hospital, The Tweed Hospital, Wagga Wagga Base Hospital, Wollongong Hospital)  3, PAEDIATRIC MAJOR TRAUMA SERVICE (Sydney Children's Hospital, Children's Hospital Westmead, John Hunter Children's Hospital)  4, RURAL LOCAL HEALTH (all other rural hospitals)  5, METROPOLITAN LOCAL HEALTH (all other metropolitan hospitals) |
| 4.1-AS | *Click on the box that best fits your current MAIN AREA OF SURGICAL PRACTICE at this hospital (choose one only)* | 1, General Surgery  2, Rural Surgery  3, Trauma Surgery  4, Acute Care Surgery  5, Vascular Surgery  6, Upper Gastrointestinal Surgery  7, Lower Gastrointestinal Surgery  8, Breast/Endocrine Surgery  9, Head and Neck Surgery  12, Paediatric Surgery  11, Research  10, Other |
| 4.2-AS | *Please name your current MAIN AREA OF SURGICAL PRACTICE not listed above* | Free text |
|  |  | Survey continues next page |

| 5.1-AS | *Indicate any OTHER OR SECONDARY AREAS OF SURGICAL INTEREST you have at this hospital. (please click on as many as apply)* | 1, Adult General Surgery  2, Rural Surgery  3, Trauma Surgery  4, Acute Care Surgery  5, Vascular Surgery  6, Upper Gastrointestinal Surgery  7, Lower Gastrointestinal Surgery  8, Breast/endocrine Surgery  9, Head and Neck Surgery  10, Paediatric Surgery  13, Research  12, None  11, Other |
| --- | --- | --- |
| 5.2-AS | *Please list OTHER OR SECONDARY AREAS OF SURGICAL PRACTICE not listed above* | Free text |
| 4.1-PS | *Click on the box that best fits your CURRENT MAIN AREA OF PAEDIATRIC SURGICAL PRACTICE at this hospital (choose one only)* | 1, General Paediatric Surgery  2, Trauma Surgery  3, Acute Care Surgery  4, Upper Gastrointestinal Surgery  5, Lower Gastrointestinal Surgery  6, Head and Neck Surgery  7, Oncology Surgery  8, Hepatobiliary Surgery  9, Urology  10, Neonatal Surgery  12, Thoracic Surgery  13, Research  11, Other |
| 4.2-PS | *Please name CURRENT MAIN AREA OF PAEDIATRIC SURGICAL PRACTICE not listed above.* | Free text |
| 5.1-PS | *Indicate any SECONDARY OR OTHER AREAS OF PAEDIATRIC SURGICAL INTEREST you have at this hospital. (please click on as many as apply)* | 1, General Paediatric Surgery  2, Trauma Surgery  3, Acute Care Surgery  4, Upper Gastrointestinal Surgery  5, Lower Gastrointestinal Surgery  6, Head and Neck Surgery  7, Oncology Surgery  8, Hepatobiliary Surgery  9, Urology  10, Neonatal Surgery  12, Thoracic Surgery  13, Research  11, Other |
| 5.2-PS | *Please name any other SECONDARY OR OTHER AREAS OF PAEDIATRIC SURGICAL INTEREST not listed above* | Free text |
| Survey continues next page | | |

| The following group of questions is about your surgical training and other qualifications | | |
| --- | --- | --- |
| 6-AS | *In what year were you awarded your FRACS in Adult General Surgery?* | Free text |
| 6-PS | *In what year were you awarded your FRACS in Paediatric Surgery?* | Free text |
| 7.1 | Do you have other post-graduate formal educational or surgical sub-specialty qualifications? | 1, Yes  0, No |
| 7.2 | *Please list any other post graduate formal educational or surgical sub-specialty qualifications* | Free text |
| 8.1 | Please indicate any TRAUMA RELATED EDUCATION, QUALIFICATIONS, TRAINING or COURSES you have undertaken. (check all that apply) | 1, Emergency Management of Severe Trauma (EMST)  2, Advanced Paediatric Life Support (APLS)  3, Definitive Surgical Trauma Course (DSTC)  4, Emergency Trauma Management (ETM)  5, FAST emergency ultrasound course  6, Tiny Tots trauma  7, International Trauma Life Support (ITLS)  14, Care of the Critically Ill Surgical Patient (CCriSP)  15, MIMMS Basic  16, MIMMS Advanced  8, Formal post FRACS trauma fellowship in Australia  9, Formal post FRACS trauma fellowship overseas  10, Masters degree in trauma or related field  11, PhD in trauma or related field  12, Fellowship training experience PTC  13, Fellowship training experience RTC  14, Fellowship training experience ATC  15, None  16, Other |
| 8.2 | *List other trauma related qualifications, courses or training you have undertaken* |  |
|  |  | Survey continues next page |

| The next group of questions relates to the management of children in your practice. | | |
| --- | --- | --- |
| 9.1.1 | Do you believe you have the skills and capacity to look after at least some ELECTIVE GENERAL SURGICAL CONDITIONS IN CHILDREN in your practice? | 1, Yes  0, No |
| 9.1.2 | *DOWN TO WHAT AGE are you comfortable looking after these elective general surgical conditions in children (in years) ?* | Free text |
| 9.1.3 | *Within the confines of the age limits above, indicate by moving the slider, the COMPLEXITY of the elective surgery you are comfortable managing in children.* | 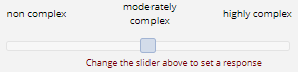 |
| 9.2.1 | Do you believe you have the skills and capacity to look after at least some EMERGENCY GENERAL SURGICAL CONDITIONS IN CHILDREN in your practice? | 1, Yes  0, No |
| 9.2.2 | *DOWN TO WHAT AGE are you comfortable looking after these emergency general surgical conditions in children (in years) ?* | Free text |
| 9.2.3 | *Within the confines of the age limits above, indicate by moving the slider, the COMPLEXITY of the emergency surgery you are comfortable managing in children.* | 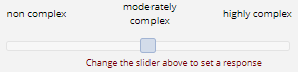 |
| 9.3.1 | Do you believe you have the skills and capacity to look after at least some INJURED CHILDREN in your practice? | 1, Yes  0, No |
| 9.3.2 | *DOWN TO WHAT AGE are you comfortable looking after these injured children (in years)?* | Free text |
| 9.3.3 | *Within the confines of the age limits above, indicate by moving the slider, the COMPLEXITY of the injury in children you are comfortable managing.* | 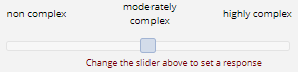 |
| 9.4 | Please feel free to add anything or clarify any of the above responses about the management of children in your practice. | Free text |
|  |  | Survey continues next page |

| The next group of questions are specifically about SPLENIC INJURY | | |
| --- | --- | --- |
| 10.1 | Approximately how many patients AGE 0-8 with SPLENIC INJURY have you managed in the most recent 12 months? | Free text |
| 10.2 | Approximately how many patients AGE 9-16 with SPLENIC INJURY have you managed in the most recent 12 months? | Free text |
| 10.3 | Approximately how many patients AGE 17-25 with SPLENIC INJURY have you managed in the most recent 12 months? | Free text |
| 10.4 | Please feel free to clarify your answers | Free text |
| 11.1-11.6 PTC | *With regards to children and young people with ISOLATED SPLENIC INJURY presenting to your hospital, while you are on call, what is your preference for their immediate care?*   (a) *0-4; (b) 5-8; (c) 9-12;*   *(d) 13-16; (e) 17-19; (f) 20-25* | 1, Keep under my care  2, Transfer to another surgeon at my hospital  3, Transfer to Regional Trauma Service  4, Transfer to Adult Major Trauma Service  6, Transfer to Metropolitan Local Health Service |
| 11.1-11.6 ATC | *With regards to children and young people with ISOLATED SPLENIC INJURY presenting to your hospital, while you are on call, what is your preference for their immediate care?*   (a) *0-4; (b) 5-8; (c) 9-12;* (b) *(d) 13-16; (e) 17-19; (f) 20-25* | 1, Keep under my care  2, Transfer to another surgeon at my hospital  3, Transfer to Regional Trauma Service  5, Transfer to Paediatric Major Trauma Service |
| 11.1-11.6 RTC | *With regards to children and young people with ISOLATED SPLENIC INJURY presenting to your hospital, while you are on call, what is your preference for their immediate care?*   (a) *0-4; (b) 5-8; (c) 9-12;*   *(d) 13-16; (e) 17-19; (f) 20-25* | 1, Keep under my care  2, Transfer to another surgeon at my hospital  4, Transfer to Adult Major Trauma Service  5, Transfer to Paediatric Major Trauma Service |
| 11.1-11.6 MLH | *With regards to children and young people with ISOLATED SPLENIC INJURY presenting to your hospital, while you are on call, what is your preference for their immediate care?*   (a) *0-4; (b) 5-8; (c) 9-12;* (b) *(d) 13-16; (e) 17-19; (f) 20-25* | 1, Keep under my care  2, Transfer to another surgeon at my hospital  3, Transfer to Regional Trauma Service  4, Transfer to Adult Major Trauma Service  5, Transfer to Paediatric Major Trauma Service |
| 11.1-11.6 RLH | *With regards to children and young people with ISOLATED SPLENIC INJURY presenting to your hospital, while you are on call, what is your preference for their immediate care?*   (a) *0-4; (b) 5-8; (c) 9-12;* (b) *(d) 13-16; (e) 17-19; (f) 20-2525* | 1, Keep under my care  2, Transfer to another surgeon at my hospital  3, Transfer to Regional Trauma Service  4, Transfer to Adult Major Trauma Service  5, Transfer to Paediatric Major Trauma Service |
|  |  | Survey continues next page |

| The following questions ask about the use of guidelines and participation in trauma audit | | |
| --- | --- | --- |
| 12.1 | Are you familiar with published guidelines for the management of splenic injury in ADULTS? | 1, unfamiliar  2, somewhat unfamiliar  3, somewhat familiar  4, familiar |
| 12.2 | Do you use guidelines for the management of splenic injury in ADULTS? | 1, never use  2, rarely use  3, occasionally use  4, often use  5, almost always use  6, always use |
| 12.3 | Do you believe the use of management guidelines has the potential to improve the outcome of splenic injury in YOUNG ADULTS presenting to your institution? | 1, strongly disagree  2, moderately disagree  3, mildly disagree  4, mildly agree  5, moderately agree  6, strongly agree |
| 12.4 | Are you involved in formal audit of ADULT trauma management outcomes at your institution? | 1, yes  2, no |
| 12.5 | Are you familiar with published guidelines for the management of splenic injury in CHILDREN? | 1, unfamiliar  2, somewhat unfamiliar  3, somewhat familiar  4, familiar |
| 12.6 | Do you use guidelines for the management of splenic injury in CHILDREN? | 1, never use  2, rarely use  3, occasionally use  4, often use  5, almost always use  6, always use |
| 12.7 | Do you believe the use of management guidelines has the potential to improve the outcome of splenic injury in CHILDREN presenting to your institution? | 1, strongly disagree  2, moderately disagree  3, mildly disagree  4, mildly agree  5, moderately agree  6, strongly agree |
| 12.8 | Are you involved in formal audit of PAEDIATRIC trauma management outcomes at your institution? | 1, yes  2, no |
|  |  | Survey continues next page |

| The next two questions are about the availability of personnel and resources for trauma care at your hospital. | | |
| --- | --- | --- |
| 13.1-13-6 | Do you believe that your hospital has the mix of APPROPRIATELY SKILLED PERSONNEL that can be relied upon to care for the following age groups with splenic injury?   (a) 0-4 (b) 5-8 (c) 9-12 (d) 13-16 (e) 17-19 (f) 20-25 | 1, Strongly disagree  2, Moderately disagree  3, Mildly disagree  4, Mildly agree  5, Moderately agree  6, Strongly agree |
| 13.7 | Please feel free to comment or clarify if you wish | Free text |
| 14.1-14.6 | Do you believe that your hospital has the PHYSICAL RESOURCES required to care for the following age groups with splenic injury?   (a) 0-4 (b) 5-8 (c) 9-12 (d) 13-16 (e) 17-19 (f) 20-25 | 1, Strongly disagree  2, Moderately disagree  3, Mildly disagree  4, Mildly agree  5, Moderately agree  6, Strongly agree |
| 14.7 | Please feel free to comment or clarify if you wish | Free text |
|  |  | Survey continues next page |

| Clinical Scenarios | | |
| --- | --- | --- |
| For the following clinical scenarios involving SPLENIC INJURY, please indicate the management strategy you are most likely to pursue (AFTER APPROPRIATE RESUSCITATION).  Options are  OM - (OPERATIVE MANAGEMENT) - very likely that you will initially pursue OPERATIVE MANAGEMENT.  NOM - (NON-OPERATIVE MANAGEMENT) -very likely that you will initially pursue NON-OPERATIVE MANAGEMENT.  AE - (ANGIO-EMBOLISATION) - very likely that you will initially pursue ANGIO-EMBOLISATION.  TRANSFER - very likely that you will transfer the patient.  Please note, although an answer is required for each scenario, there is room to clarify your response if you wish. | | |
| 15.1 | Grade IV isolated splenic injury,  Haemodynamically STABLE on admission.  Moderate intraperitoneal blood, with NO CONTRAST EXTRAVASATION (‘blush’) on abdominal CT-scan.   (a) 10-year-old   18-year-old | 1, OM  2, NOM  3, AE  4, TRANSFER |
| 15.1.1 | Please feel free to add anything or clarify your answer. (for example, hospital location, personnel or resource factors, or other patient factors that may influence your decision) | Free text |
| 15.2 | Grade IV isolated splenic injury.  Haemodynamically STABLE on admission.  Moderate intraperitoneal blood, with CONTRAST EXTRAVASATION (‘blush’) PRESENT on abdominal CT-scan.   (a) 10-year-old   18-year-old | 1, OM  2, NOM  3, AE  4, TRANSFER |
| 15.2.1 | Please feel free to add anything or clarify your answer. (for example, hospital location, personnel or resource factors, or other patient factors that may influence your decision) | Free text |
| 15.3 | Grade IV isolated splenic injury.  Surgeon suspects intra-abdominal bleeding on admission.  Sustained response to  10mls/kg of normal saline and transfusion of 10mls/kg packed red blood cells (10-year-old)  Saline bolus and transfusion of 2 units packed red blood cells (18-year-old).   (a) 10-year-old   18-year-old | 1, OM  2, NOM  3, AE  4, TRANSFER |
| 15.3.1 | Please feel free to add anything or clarify your answer. (for example, hospital location, personnel or resource factors, or other patient factors that may influence your decision) | Free text |
| 15.4 | Grade IV splenic injury WITH GRADE II LIVER LACERATION.  Haemodynamically STABLE on admission.  Moderate intraperitoneal blood, with NO CONTRAST EXTRAVASATION (‘blush’) on abdominal CT-scan.   (a) 10-year-old   18-year-old | 1, OM  2, NOM  3, AE  4, TRANSFER |
|  |  | Survey continues next page |

| 15.4.1 | Please feel free to add anything or clarify your answer. (for example, hospital location, personnel or resource factors, or other patient factors that may influence your decision) | Free text |
| --- | --- | --- |
| 15.5 | Grade IV splenic injury with grade II liver laceration.  Haemodynamically STABLE on admission.  Moderate intraperitoneal blood, with CONTRAST EXTRAVASATION (‘blush’) PRESENT on abdominal CT-scan (from the spleen).   (a) 10-year-old   18-year-old | 1, OM  2, NOM  3, AE  4, TRANSFER |
| 15.5.1 | Please feel free to add anything or clarify your answer. (for example, hospital location, personnel or resource factors, or other patient factors that may influence your decision) | Free text |
| 15.6 | Grade IV splenic injury with grade II liver laceration.  Surgeon suspects intra-abdominal bleeding on admission.  Initial response to  10mls/kg normal saline and transfusion of 10mls/kg packed red blood cells (10-year-old)  Saline bolus plus transfusion of 2 units packed red blood cells (18-year-old).  Develops recurrent haemodynamic instability at 4 hours which responds to further appropriate blood product administration.  Moderate intraperitoneal blood, with CONTRAST EXTRAVASATION (‘blush’) PRESENT on abdominal CT-scan (from the spleen).   (a) 10-year-old   18-year-old | 1, OM  2, NOM  3, AE  4, TRANSFER |
| 15.6.1 | Please feel free to add anything or clarify your answer. (for example, hospital location, personnel or resource factors, or other patient factors that may influence your decision) | Free text |
| 15.7 | Grade IV splenic injury, INTUBATED AND VENTILATED.  SMALL EXTRADURAL HAEMATOMA on head CT-scan.  Haemodynamically STABLE on admission.  Moderate intraperitoneal blood, with NO CONTRAST EXTRAVASATION (‘blush’) on abdominal CT-scan.   (a) 10-year-old   18-year-old | 1, OM  2, NOM  3, AE  4, TRANSFER |
|  |  | Survey continues next page |

| 15.7.1 | Please feel free to add anything or clarify your answer. (for example, hospital location, personnel or resource factors, or other patient factors that may influence your decision) | Free text |
| --- | --- | --- |
| 15.8 | Grade IV splenic injury, INTUBATED AND VENTILATED.  SMALL EXTRADURAL HAEMATOMA on head CT-scan.  Surgeon suspects intra-abdominal bleeding on admission.  Sustained response to  10mls/kg normal saline and transfusion of 10mls/kg packed red blood cells (10-year-old)  Saline bolus and transfusion of 2 units packed cells (18-year-old).  Moderate intraperitoneal blood, with NO CONTRAST EXTRAVASATION (‘blush’) on abdominal CT-scan.   (a) 10-year-old   18-year-old | 1, OM  2, NOM  3, AE  4, TRANSFER |
| 15.8.1 | Please feel free to add anything or clarify your answer. (for example, hospital location, personnel or resource factors, or other patient factors that may influence your decision) | Free text |
| 15.9 | Grade IV splenic injury. INTUBATED AND VENTILATED.  SMALL EXTRADURAL HAEMATOMA on head CT-scan.  Surgeon suspects intra-abdominal bleeding on admission.  Responds initially to 10mls/kg normal saline and transfusion of 10mls/kg packed red blood cells (10-year-old)  Saline bolus and transfusion of 2 units packed cells (18-year-old)  Develops recurrent haemodynamic instability at 4 hours which responds to further appropriate blood product administration.  Moderate intraperitoneal blood, and CONTRAST EXTRAVASATION (‘blush’) PRESENT on abdominal CT-scan.   (a) 10-year-old   18-year-old | 1, OM  2, NOM  3, AE  4, TRANSFER |
| 15.9.1 | Please feel free to add anything or clarify your answer. (for example, hospital location, personnel or resource factors, or other patient factors that may influence your decision) | Free text |
|  |  | Survey continues next page |

| 15.10 | Grade IV splenic injury, grade II liver injury.  INTUBATED AND VENTILATED.  SMALL EXTRADURAL HAEMATOMA on head CT-scan.  Surgeon suspects intra-abdominal bleeding on admission.  Responds initially to 10mls/kg saline and transfusion of 10mls/kg packed red blood cells (10-year-old)  Saline bolus and transfusion of 2 units packed red blood cells (18-year-old).  Develops recurrent haemodynamic instability at 4 hours, which transiently responds to further appropriate blood product administration.  Haemodynamic instability recurs over the following 2 hours, which is, again, appropriately resuscitated.  Moderate intraperitoneal blood, and NO CONTRAST EXTRAVASATION (‘blush’) on abdominal CT-scan.   (a) 10-year-old   18-year-old | 1, OM  2, NOM  3, AE  4, TRANSFER |
| --- | --- | --- |
| 15.10.1 | Please feel free to add anything or clarify your answer. (for example, hospital location, personnel or resource factors, or other patient factors that may influence your decision) | Free text |
| Background information | | |
| 16 | What is your age in years? | Free text |
| 17 | What is your gender? | 1, Male  2, Female  3, Other  4, Prefer not to say |
| 18 | Where did you attend medical school? | 1, New South Wales  2, Queensland  3, South Australia  4, Victoria  5, Western Australia  6, Tasmania  7, Australian Capital Territory  8, Overseas |
| 18.1 | *In which country did you attend Medical School?* | Free text |
| 19 | IN what year did you graduate from medical school? | Free text |
| 20 | Please feel free to add any other comments about the management of splenic injury in children and young adults at your hospital and/or in the NSW Trauma System |  |
| The survey is now complete. Thank you very much for your time. | | |
| 21 | Please indicate if you would like to receive a copy of the survey results | 1, Yes  2, No |
| 21.1 | *Please provide your email address to receive survey results* | Free text |

| Supplementary Table 3a: Responses for scenarios 1, 4, 7 and 8 by hospital category  All grade IV splenic injury with moderate intraperitoneal blood and no contrast blush on abdominal CT-scan. Pediatric guideline: ATOMAC = the North American Paediatric Trauma Consortium (Arizona-Texas-Oklahoma-Memphis-Arkansas). Adult guideline: WTA = Western Trauma Association. SI = splenic injury. HD=haemodynamic. IA = intra-abdominal. IP = intraperitoneal. NS=normal saline, PRBC=packed red blood cells. OM = operative management. NOM = non-operative management. AE =angioembolisation. PTC = paediatric trauma centre. ATC = metropolitan adult trauma centre. RTC = regional/rural trauma centre. MLH = metropolitan local hospital. RLH = regional/rural local hospital. OR = odds ratio. Unif = uniform. Transfer and NOM analysed as one variable for treatment choice and alignment with guidelines. * denotes each paediatric surgeon at an adult centre. | | | | | | | | | | | | |
| --- | --- | --- | --- | --- | --- | --- | --- | --- | --- | --- | --- | --- |
| Scenario 1 | Grade IV isolated SI. HD stable on admission.  CT abdomen– Moderate IP blood. No contrast blush.  Pediatric – NOM/TRANSFER. Adult – NOM/AE/TRANSFER | | | | | | | | | | | |
| number | PTC N=12(%) | | ATC N=16(%) | | RTC N=25(%)** | | MLH N=6(%)* | | RLH N=9(%) | | p | |
| age | 10 | 18 | 10 | 18 | 10 | 18 | 10 | 18 | 10 | 18 | 10 | 18 |
| OM |  |  |  |  | 1(4) | 1(4) |  |  |  |  |  |  |
| NOM | 12 (100) | 10(83) | 7(44) | 16(100) | 14**(56) | 23**(92) | 3*(50) | 3(50) | 2(22) | 7(78) | 0.32 | 0.64 |
| AE |  |  |  |  |  | 1(8) | 1(17) | 1(17) |  |  |  |  |
| TF |  | 2(17) | 9(56) |  | 10(40) |  | 2(33) | 2*((33) | 7(78) | 2(22) |  |  |
| C/W ATOMAC | 12(100) | 12(100) | 16(100) | 16(100) | 24(96) | 23**(92) | 5(83) | 5*(83) | 9(100) | 9(100) | 0.38 | 0.40 |
| C/W WTA | 12(100) | 12(100) | 16(100) | 16(100) | 24(96) | 24(96) | 6(100) | 6(100) | 9(100) | 9(100) | 1.00 | 1.00 |
| Scenario 4 - | Grade IV SI with grade II liver laceration. HD stable on admission.  CT abdomen - Moderate IP blood. No contrast blush.  ATOMAC – NOM/TRANSFER. WTA – NOM/AE/TRANSFER. | | | | | | | | | | | |
| number | PTC N=12(%) | | ATC N=14(%) | | RTC N=23(%)* | | MLH N=5(%)* | | RLH N=9(%) | | p | |
| age | 10 | 18 | 10 | 18 | 10 | 18 | 10 | 18 | 10 | 18 | 10 | 18 |
| OM |  |  |  |  |  |  |  |  |  |  |  |  |
| NOM | 12(100) | 11(92) | 5(36) | 12(86) | 9(39) | 18(78) | 3*(60) | 2(40) | 3(33) | 8(89) | uniform | 0.54 |
| AE |  |  |  | 2(14) |  | 1(4) |  |  |  |  |  |  |
| TF |  | 1(8) | 9(64) |  | 14*(61) | 4*(17) | 2(40) | 3*(60) | 6(67) | 1(11) |  |  |
| C/W ATOMAC | 12(100) | 12(100) | 14(100) | 12(86) | 23*(100) | 22(96) | 5(100) | 5(100) | 9(100) | 9(100) | uniform | 0.54 |
| C/W WTA | 12(100) | 12(100) | 14(100) | 14(100) | 23(100) | 23(100) | 5(100) | 5(100) | 9(100) | 9(100) | uniform | uniform |
| Table continues next page | | | | | | | | | | | | |

### Scenario responses and mapping to guidelines

| Scenario 7 | Grade IV SI. Intubated and ventilated. HD stable on admission. Small extradural haematoma on head CT.  CT abdomen – Moderate IP blood. No contrast blush.  ATOMAC – NOM/TRANSFER. WTA – NOM/AE/TRANSFER. | | | | | | | | | | | |
| --- | --- | --- | --- | --- | --- | --- | --- | --- | --- | --- | --- | --- |
| number | PTC N=12(%) | | ATC N=14(%) | | RTC N=21(%)* | | MLH N=5(%)* | | RLH N=9(%) | | p | |
| age | 10 | 18 | 10 | 18 | 10 | 18 | 10 | 18 | 10 | 18 | 10 | 18 |
| OM |  |  |  |  |  |  |  |  |  | 1(11) |  |  |
| NOM | 12(100) | 10(83) | 5(36) | 12(86) | 4(19) | 8(38) |  |  |  |  | 0.23 | 0.10 |
| AE |  |  |  | 2(16) |  |  |  |  | 1(11) |  |  |  |
| TF |  | 2(17) | 9 (64) |  | 17*(81) | 13*(62) | 5*(100) | 5*(100) | 8(89) | 8(89) |  |  |
| C/W ATOMAC | 12(100) | 12(100) | 14(100) | 12(86) | 14*(100) | 21*(100) | 5*(100) | 5*(100) | 8(89) | 8(89) | 0.20 | 0.18 |
| C/W WTA | 12(100) | 12(100) | 12(100) | 14(100) | 21(100) | 21(100) | 5(100) | 5(100) | 9(100) | 8(89) | uniform | uniform |
| Scenario 8 | Grade IV SI. Intubated and ventilated. Surgeon suspects IA bleeding. Small extradural haematoma on head CT.  Sustained response to 10mls/kg NS + 10mls/kg PRBCs (age 10)/NS bolus +1 Unit PRBCs (age 18).  CT abdomen – Moderate IP blood. No contrast blush.  ATOMAC – NOM/TRANSFER. WTA – AE/TRANSFER. | | | | | | | | | | | |
| number | PTC N=12(%) | | ATC N=14(%) | | RTC N=21(%)* | | MLH N=5(%)* | | RLH N=9(%) | | p | |
| age | 10 | 18 | 10 | 18 | 10 | 18 | 10 | 18 | 10 | 18 | 10 | 18 |
| OM |  |  |  |  |  | 1(5) |  |  |  | 1(11) |  |  |
| NOM | 11 (92) | 10(83) | 4(29) | 13(93) | 4(19) | 7(33) |  |  |  |  | 0.84 | 0.86 |
| AE | 1 (8) |  |  | 1(7) | 1(5) | 2(10) |  |  |  |  |  |  |
| TF |  | 2(16) | 10(71) |  | 16*(76) | 11*(52) | 5*(100) | 5*(100) | 9(100) | 8(89) |  |  |
| C/W ATOMAC | 11(92) | 12(100) | 14(100) | 13(93) | 20*(95) | 18*(86) | 5*(100) | 5*(100) | 9(100) | 8(89) | 0.94 | 0.71 |
| C/W WTA | 1(8) | 2(17) | 10(71) | 1(7) | 17(81) | 13(62) | 5(100) | 5(100) | 9(100) | 8(100) | **<0.001** | **<0.001** |

| Supplementary Table 3b: Responses for scenarios 1, 4, 7 and 8 by surgeon type  All grade IV splenic injury with moderate intraperitoneal blood and no contrast blush on abdominal CT-scan. Pediatric guideline: ATOMAC = the North American Paediatric Trauma Consortium (Arizona-Texas-Oklahoma-Memphis-Arkansas). Adult guideline: WTA = Western Trauma Association. SI = splenic injury. HD=haemodynamic. IA = intraabdominal. IP = intra-peritoneal. NS=normal saline, PRBC=packed red blood cells. OM = operative management. NOM = non-operative management. AE = angioembolisation. AS =adult surgeon. PS = paediatric surgeon. M = metropolitan. R = regional/rural. OR = odds ratio. Unif = uniform. Transfer and NOM analysed as one variable for treatment choice and alignment with guidelines. | | | | | | | | | | | | | | |
| --- | --- | --- | --- | --- | --- | --- | --- | --- | --- | --- | --- | --- | --- | --- |
| Scenario 1 | Grade IV isolated SI. HD stable on admission.  CT abdomen– Moderate IP blood. No contrast blush.  ATOMAC = NOM/TRANSFER. WTA = NOM/AE/TRANSFER. | | | | | | | | | | | | | |
| number | PS N=15(%) | | AS N=53(%) | | p – PS vs AS | | M-AS N=21(%) | | R-AS N=32(%) | | p – M-AS vs R-AS | | p - PS 10 vs 18 | p - AS 10 vs 18 |
| age | 10 | 18 | 10 | 18 | 10 | 18 | 10 | 18 | 10 | 18 | 10 | 18 |  |  |
| OM |  |  | 1(2) | 1(2) |  |  |  |  | 1(3) | 1(3) |  |  |  |  |
| NOM | 15(100) | 12(80) | 23(43) | 47(89) | 1.00 | 1.00 | 9(43) | 19(90) | 14((44) | 28(87) | 0.64 | 1.00 | uniform | 1.00 |
| AE |  |  | 1(2) | 2(4) |  |  | 1(5) | 1(5) |  | 1(3) |  |  |  |  |
| TF |  | 3 (20) | 28(53) | 3(6) |  |  | 1(952) | 1(5) | 17(53) | 2(6) |  |  |  |  |
| C/W ATOMAC | 15(100) | 15(100) | 51(96) | 50(94) | 1.00 | 1.00 | 20(95) | 20(95) | 31(97) | 30(94) | 1.00 | 1.00 | uniform | 1.00 |
| C/W WTA | 15(100) | 15(100) | 52(98) | 52 (98) | 1.00 | 1.00 | 21(100) | 21(100) | 31(97) | 31(97) | 1.00 | 1.00 | uniform | 1.00 |
| Scenario 4 - | Grade IV SI with grade II liver laceration. HD stable on admission.  CT abdomen - Moderate IP blood. No contrast blush.  ATOMAC = NOM/TRANSFER. WTA = NOM/AE/TRANSFER. | | | | | | | | | | | | | |
| number | PS N=14(%) | | AS N=49(%) | | p – PS vs AS | | M-AS N=18(%) | | R-AS N=31(%) | | p – M-AS | | p - PS 10 vs 18 | p - AS 10 vs 18 |
| age | 10 | 18 | 10 | 18 | 10 | 18 | 10 | 18 | 10 | 18 | 10 | 18 |  |  |
| OM |  |  |  |  |  |  |  |  |  |  |  | 0.55 | uniform | 0.24 NS |
| NOM | 13(93) | 11(79) | 19(39) | 40(82) | uniform | 1.00 | 7(39) | 14(78) | 12(39) | 26(84) | uniform |  |  |  |
| AE |  |  |  | 3(6) |  |  |  | 2(11) |  | 1(3) |  |  |  |  |
| TF | 1(7) | 3(21) | 30(61) | 6(12) |  |  | 11(61) | 2(11) | 19(61) | 4(13) |  |  |  |  |
| C/W ATOMAC | 14(100) | 14(100) | 49(100) | 46(94) | uniform | 1.00 | 18(100) | 16(89) | 31(100) | 30(97) | uniform | 0.55 | uniform | 0.24 |
| C/W WTA | 14(100) | 14(100) | 49(100) | 49(100) | uniform | uniform |  |  |  |  | uniform | uniform | uniform | uniform |
| Table continues next page | | | | | | | | | | | | | | |

| Scenario 7 | Grade IV SI. Intubated and ventilated. HD stable on admission. Small extradural haematoma on head CT.  CT abdomen – Moderate IP blood. No contrast blush.  ATOMAC = NOM/TRANSFER. WTA = NOM/AE/TRANSFER. | | | | | | | | | | | | | |
| --- | --- | --- | --- | --- | --- | --- | --- | --- | --- | --- | --- | --- | --- | --- |
| number | PS N=14(%) | | AS N=47(%) | | p – PS vs AS | | M-AS N= 18(%) | | R-AS N = 29(%) | | p – M-AS vs R-AS | | p - PS 10 vs 18 | p - AS 10 vs 18 |
| age | 10 | 18 | 10 | 18 | 10 | 18 | 10 | 18 | 10 | 18 | 10 | 18 |  |  |
| OM |  |  |  | 1(2) |  |  |  |  |  | 1(3) | 1.00 | 0.14 | uniform | 0.62 |
| NOM | 12(86) | 10(71) | 9(19) | 20(43) | 1.00 | 1.00 | 5(28) | 12(67) | 4(14) | 8(28) |  |  |  |  |
| AE |  |  | 1(2) | 2(4) |  |  |  | 2(11) | 1(3) |  |  |  |  |  |
| TF | 2(14) | 4(29) | 37(79) | 24(51) |  |  | 13(72) | 4(22) | 24(83) | 20(69) |  |  |  |  |
| C/W ATOMAC | 14(100) | 14(100) | 46(98) | 44(94) | 1.00 | 1.00 | 18 (100) | 16(89) | 28(97) | 28(97) | 1.00 | 0.55 | uniform | 0.62 |
| C/W WTA | 14(100) | 14(100) | 47(100) | 46(98) | 1.00 | 1.00 | 18(100) | 18(100) | 29(100) | 28(97) | Uniform | 1.00 | uniform | 0.61 |
| Scenario 8 | Grade IV SI. Intubated and ventilated. Surgeon suspects IA bleeding. Small extradural haematoma on head CT.  Sustained response to 10mls/kg NS + 10mls/kg PCs (child)/NS bolus +1 Unit PCs (YA)  CT abdomen – Moderate IP blood. No contrast blush.  ATOMAC – NOM/TRANSFER. WTA – AE/TRANSFER. | | | | | | | | | | | | | |
| number | PS N=14(%) | | AS N=47(%) | | p – PS vs AS, OR (95%CI) | | M-AS N=18(%) | | R-AS N=29(%) | | p – M-AS vs R-AS OR (95%CI) | | p - PS 10 vs 18 | p - AS 10 vs 18 |
| age | 10 | 18 | 10 | 18 | 10 | 18 | 10 | 18 | 10 | 18 | 10 | 18 |  |  |
| OM |  |  |  | 2(4) |  |  |  |  |  | 1(3) | 1 NS | 0.78 NS | 1 NS | 0.23 NS |
| NOM | 11(79) | 10(71) | 8(17) | 20(43) | 0.41 | 1 NS | 4(22) |  | 4(14) | 8(28) |  |  |  |  |
| AE | 1(7) |  | 1(2) | 3(6) |  |  |  | 1(6) | 1(3) | 2(7) |  |  |  |  |
| TF | 2(14) | 4(29) | 38(81) | 22(47) |  |  | 14(78) | 1(6) | 24(83) | 18(62) |  |  |  |  |
| C/W ATOMAC | 13(93) | 14(100) | 46(98) | 42(89) | 0.40  OR 0.29  (0.003-23.88) | 0.58 NS | 18(100) | 17(94) | 28(97) | 25(86) | 1 NS  OR 0  (0-62.78) | 0.63 NS  OR 0.37  (0.007-4.23) | 1 NS  OR inf  (0.03-inf) | 0.20 NS  OR 0.18  (0.003-1.75) |
| C/W WTA | 3(21) | 4(29) | 39(83) | 25(53) | **<0.001**  **OR 0.06**  **(0.009-0.29)** | 0.13 NS  OR 0.36  (0.07-1.47) | 14(78) | 5(28) | 25(86) | 20(69) | 0.69  OR 1.76  (0.28-11.09) | **<0.05**  **OR 5.54**  **(1.35-26.48)** | 1 NS  OR 1.45  (0.19-12.44) | **<0.01**  **OR 0.24**  **(0.08-0.66)** |

| Supplementary Table 3c: Responses for scenarios 2, 3, 5, 6 and 9 by hospital category  All grade IV splenic injury with moderate intraperitoneal blood and contrast blush on abdominal CT-scan. Pediatric guideline: ATOMAC = the North American Paediatric Trauma Consortium (Arizona-Texas-Oklahoma-Memphis-Arkansas). Adult guideline: WTA = Western Trauma Association. SI = splenic injury. HD=haemodynamic. IA = intra-abdominal. IP = intra-peritoneal. NS=normal saline, PRBC=packed red blood cells. OM = operative management. NOM = non-operative management. AE = angioembolisation. PTC = paediatric trauma centre. ATC = metropolitan adult trauma centre. RTC = regional/rural trauma centre. MLH = metropolitan local hospital. RLH = regional/rural local hospital. OR = odds ratio. Unif = uniform. Transfer and NOM analysed as one variable for treatment choice and alignment with guidelines. * denotes each Paediatric Surgeon at an adult centre | | | | | | | | | | | | |
| --- | --- | --- | --- | --- | --- | --- | --- | --- | --- | --- | --- | --- |
| Scenario 2 | Grade IV isolated SI. HD stable on admission.  CT abdomen – Moderate IP blood. Contrast blush.  ATOMAC- NOM/TRANSFER. WTA - AE/TRANSFER. | | | | | | | | | | | |
| number | PTC N=12(%) | | ATC N=15(%) | | RTC N=25(%)** | | MLH N=6(%)* | | RLH N=9(%) | | p | |
| age | 10 | 18 | 10 | 18 | 10 | 18 | 10 | 18 | 10 | 18 | 10 | 18 |
| OM |  |  |  | 1(7) | 1(4) | 4(16) |  |  |  |  |  |  |
| NOM | 9(75) | 6(50) | 3(20) | 2(13) | 7**(28) | 7*(28) | 1*(17) |  |  | 1(11) | 0.89 NS | 0.06 |
| AE | 3(25) | 4(33) | 4(27) | 12(80) | 9 (36) | 12 (48) | 3(50) | 4(67) | 2(22) | 3(33) |  |  |
| TF |  | 2(17) | 8(53) |  | 8(32) | 2*(8) | 2(33) | 2*(33) | 7(78) | 5(56) |  |  |
| C/W ATOMAC | 9(75) | 8(67) | 11(73) | 2(13) | 15**(60) | 9**(36) | 3*(50) | 2*(33) | 7(78) | 6(67) | 0.68 NS | **<0.05** |
| C/W WTA | 3(25) | 6(50) | 12(80) | 12(80) | 17(68) | 14(56) | 5(83) | 6(100) | 9(100) | 8(89) | **<0.01** | 0.06 |
| Scenario 3 - | Grade IV isolated SI. Surgeon suspects IA bleeding.  Sustained response to 10mls/kg NS + 10mls/kg PRBCs (age 10)/NS bolus +1 Unit PRBCs (age 18).  CT abdomen - Moderate IP blood. Contrast blush.  ATOMAC – NOM. WTA – AE/TRANSFER. | | | | | | | | | | | |
| number | PTC N=12(%) | | ATC N=14(%) | | RTC N=23(%)* | | MLH N=5(%)* | | RLH N=9(%) | | p | |
| age | 10 | 18 | 10 | 18 | 10 | 18 | 10 | 18 | 10 | 18 | 10 | 18 |
| OM |  |  |  | 1(7) | 2(9) | 5(22) | 1(20) | 3(60) | 1(11) | 1(11) |  |  |
| NOM | 9(75) | 6(50) | 2(14) | 1(7) | 4(17) | 4(17) | 1*(20) |  |  | 1(11) | 0.59 NS | **<0.01** |
| AE | 3(25) | 4(33) | 4(29) | 12(86) | 8(35) | 11(48) |  |  | 3(33) | 4(44) |  |  |
| TF |  | 2(17) | 8(57) |  | 9*(39) | 3*(13) | 3(60) | 2*(40) | 5(56) | 3(33) |  |  |
| C/W ATOMAC | 9(75) | 7(58) | 10(71) | 1(7) | 13*(56) | 6*(26) | 4*(80) | 2*(40) | 5(56) | 3(33) | 0.73 NS | 0.06 |
| C/W WTA | 3(25) | 6(50) | 12(85) | 12(85) | 17(74) | 14(61) | 3(60) | 2(40) | 8(89) | 7(78) | **<0.01** | 0.19 |
| Table continues next page | | | | | | | | | | | | |

| Scenario 5 - | Grade IV SI with grade II liver laceration. HD stable on admission.  CT abdomen - Moderate IP blood. Contrast blush.  ATOMAC – NOM/TRANSFER. WTA – AE/TRANSFER. | | | | | | | | | | | |
| --- | --- | --- | --- | --- | --- | --- | --- | --- | --- | --- | --- | --- |
| number | PTC N=12(%) | | ATC N=14(%) | | RTC N=22(%)* | | MLH N=5(%)* | | RLH N=9(%) | | p | |
| age | 10 | 18 | 10 | 18 | 10 | 18 | 10 | 18 | 10 | 18 | 10 | 18 |
| OM | 1(8) | 1(8) |  |  |  | 2(9) |  |  |  |  |  |  |
| NOM | 8(67) | 4(33) | 1(7) | 1(7) | 2(9) | 3(14) | 1*(20) |  |  | 1(11) | 0.66 NS | <0.05 |
| AE | 3(25) | 5(42) | 3(21) | 13(93) | 8(36) | 13(59) | 1(20) | 2(40) | 1(11) | 3(33) |  |  |
| TF |  | 2(17) | 10(72) |  | 12*(55) | 4*(18) | 3(60) | 3*(60) | 8(89) | 5(56) |  |  |
| C/W ATOMAC | 8(67) | 6(50) | 11(79) | 1(7) | 14*(64) | 7*(32) | 4*(80) | 3*(60) | 8(89) | 6(66.7) | 0.66 NS | <0.05 |
| C/W WTA | 3(25) | 7(58) | 13(93) | 13(93) | 20(91) | 17(77) | 4(80) | 5(100) | 9(100) | 8(89) | <0.001 | 0.19 NS |
| Scenario 6 - | Grade IV SI with grade II liver laceration. Surgeon suspects IA bleeding.  Initial response to 10mls/kg NS + 10mls/kg PRBCs (age 10)/NS bolus +1 Unit PRBCs (age 18).  Recurrent HD instability at 4 hours responsive to further blood products.  CT abdomen - Moderate IP blood. Contrast blush.  ATOMAC – NOM/AE/TRANSFER. WTA – AE/OM/TRANSFER. | | | | | | | | | | | |
| number | PTC N=12(%) | | ATC N=14(%) | | RTC N=22(%)* | | MLH N=5(%)* | | RLH N=9(%) | | p | |
| age | 10 | 18 | 10 | 18 | 10 | 18 | 10 | 18 | 10 | 18 | 10 | 18 |
| OM | 1 (8) | 2 (17) | 2(14) | 8(57) | 10(46) | 10(46) | 2(40) | 2(40) | 3(33) | 6(67) |  |  |
| NOM | 2 (16) | 2 (17) | - | - | - | - | - | - | - | 1(11) | **<0.01** | **<0.01** |
| AE | 9 (75) | 7 (58) | 2 (14) | 6(43) | 6(27) | 11*(50) | 1*(20) | 1(20) | - |  |  |  |
| TF |  | 1 (8) | 10(71) |  | 6*(27) | 1(4) | 2(40) | 2*(40) | 6(67) | 2(22) |  |  |
| C/W ATOMAC | 11(92) | 10(83) | 12(86) | 6(43) | 12*(54) | 12(54) | 3*(60) | 3*(60) | 6(67) | 3(33) | 0.11 NS | 0.16 NS |
| C/W WTA | 10(83) | 10(83) | 14(100) | 14(100) | 22(100) | 22(100) | 5(100) | 5(100) | 9(100) | 9(100) | 0.11 NS | 0.11 NS |
| Scenario 9 | Grade IV SI. Intubated and ventilated. Surgeon suspects IA bleeding. Small extradural haematoma on head CT.  Initial response to 10mls/kg NS + 10mls/kg PRBCs (age 10)/NS bolus +1 Unit PRBCs (age 18).  Recurrent HD instability at 4 hours responsive to further blood products.  CT abdomen - Moderate IP blood. Contrast blush.  ATOMAC – NOM/AE/TRANSFER. WTA – AE/OM/TRANSFER. | | | | | | | | | | | |
| number | PTC N=12(%) | | ATC N=14(%) | | RTC N=21(%)* | | MLH N=5(%)* | | RLH N=9(%) | | p | |
|  | 10 | 18 | 10 | 18 | 10 | 18 | 10 | 18 | 10 | 18 | 10 | 18 |
| OM | 1 (8) | 2 (17) | 2 (14) | 7 (50) | 9 (43) | 10(48) | 1(20) | 1(20) | 4(44) | 4(44) |  |  |
| NOM | 2 (17) | 2 (17) | 1 (7) | 1 (7) |  |  |  |  |  |  | **<0.001** | **0.01** |
| AE | 9 (75) | 6 (50) | 1 (7) | 6 (43) | 5 (24) | 7 (33.3) | 1*(20) |  |  |  |  |  |
| TF |  | 2 (17) | 10 (67) |  | 7*(33) | 4*(19) | 3(60) | 4*(80) | 5(56) | 5(56) |  |  |
| C/W ATOMAC | 11(92) | 10(83) | 12(86) | 7(50) | 12*(57) | 11*(52) | 4*(80) | 4*(80) | 5(56) | 5(56) | 0.13 NS | 0.33 NS |
| C/W WTA | 10(83) | 10(83) | 13(93) | 13(93) | 21(100) | 21(100) | 5(100) | 5(100) | 9(100) | 9(100) | 0.23 NS | 0.23 NS |

| Supplementary Table 3d: Responses for scenarios 2, 3, 5, 6 and 9 by surgeon type  All grade IV splenic injury with moderate intraperitoneal blood and contrast blush on abdominal CT-scan. Pediatric guidelines: ATOMAC = the North American Paediatric Trauma Consortium (Arizona-Texas-Oklahoma-Memphis-Arkansas). Adult guidelines: WTA = Western Trauma Association. SI = splenic injury. HD=haemodynamic. IA = intraabdominal. IP = intra-peritoneal. NS=normal saline, PRBC=packed red blood cells. OM = operative management. NOM = non-operative management. AE = angioembolisation. AS =adult surgeon. PS = paediatric surgeon. M = metropolitan. R = regional/rural. OR = odds ratio. Unif = uniform. Transfer and NOM analysed as one variable for treatment choice and alignment with guidelines. | | | | | | | | | | | | | | |
| --- | --- | --- | --- | --- | --- | --- | --- | --- | --- | --- | --- | --- | --- | --- |
| Scenario 2 | Grade IV isolated SI. HD stable on admission. CT abdomen  Moderate IP blood. Contrast blush.  ATOMAC = NOM/TRANSFER. WTA = AE/TRANSFER. | | | | | | | | | | | | | |
| number | PS N=15(%) | | AS N=52(%) | | p – PS vs AS, OR (95%CI) | | M-AS N=20(%) | | R-AS N=32(%) | | p – M-AS vs R-AS OR (95%CI) | | p - PS 10 vs 18 | p - AS 10 vs 18 |
| age | 10 | 18 | 10 | 18 | 10 | 18 | 10 | 18 | 10 | 18 | 10 | 18 |  |  |
| OM |  |  | 1(2) | 5(10) | 0.5 NS | <0.05 |  | 1(5) | 1(3) | 4(12) | 1 NS | 0.06 | 1 NS | **<0.01** |
| NOM | 12(80) | 7(47) | 8(15) | 9(17) |  |  | 3(15) | 2(10) |  | 7((22) |  |  |  |  |
| AE | 3(20) | 4(27) | 18(35) | 31(60) |  |  | 7(35) | 16(80) | 11(34) | 15(47) |  |  |  |  |
| TF |  | 4(27) | 25(48) | 7(13) |  |  | 10(50) | 1(5) |  | 6(19) |  |  |  |  |
| C/W ATOMAC | 12(80) | 11(73) | 33 (63) | 16(31) | 0.35 NS  2.28  (0.52-14.15) | **<0.01**  **OR 6.00**  **(1.49-29.92)** | 13(65) | 3(15) | 20(63) | 13(41) | 1 NS  OR 0.90  (0.23-3.30) | 0.07  OR 3.78  (0.83-24.22) | 1 NS  OR 0.70  (0.08-5.18) | **<0.01**  **OR 0.26**  **(0.10-0.62)** |
| C/W WTA | 3(20) | 8(53) | 43(88) | 38(78) | **<0.001**  **OR 0.06**  **(0.008-0.26)** | 0.2 NS  OR 0.42  (0.11-1.66) | 17(85) | 17(85) | 26(81) | 21(66) | 1 NS  OR 0.77  (0.11-4.21) | 0.20 (NS)  OR 0.34  (0.05-1.59) | 0.13 NS  OR 4.33  (0.73-34.13) | 0.34 NS  OR 0.57  (0.19-1.60) |
| Scenario 3 - | Grade IV isolated SI. Surgeon suspects IA bleeding.  Sustained response to 10mls/kg NS + 10mls/kg PRBCs (age 10)/NS bolus +1 Unit PRBCs (age 18).  CT abdomen - Moderate IP blood. Contrast blush.  ATOMAC = NOM/TRANSFER. WTA = AE/TRANSFER. | | | | | | | | | | | | | |
| number | PS N=14(%) | | AS N=49(%) | | p – PS vs AS, OR (95%CI) | | M-AS N=18(%) | | R-AS N=31(%) | | p – M-AS vs R-AS OR (95%CI) | | p - PS 10 vs 18 | p - AS 10 vs 18 |
| age | 10 | 18 | 10 | 18 | 10 | 18 | 10 | 18 | 10 | 18 | 10 | 18 |  |  |
| OM |  |  | 4(8) | 10(20) |  |  | 1(6) | 4(22) | 3(10) | 6(19) | 0.52 NS | 0.31 NS | 1 NS | **0.001** |
| NOM | 10(71) | 6(43) | 6(12) | 6(12) | 0.59 NS | <0.01 | 2(11) | 1(6) | 4((13) | 5(16) |  |  |  |  |
| AE | 3(21) | 4(29) | 15(31) | 27(55) |  |  | 4(22) | 12(67) | 11(35) | 15(48) |  |  |  |  |
| TF | 1((7) | 4(29) | 24(62) | 6(12) |  |  | 11(61) | 1(6) | 13(42) | 5(16) |  |  |  |  |
| C/W ATOMAC | 11(79) | 9(64) | 30(61) | 10(20) | 0.34  OR 2.29  (0.51-14.45) | **<0.01**  **OR 6.75**  **(1.61-32.01)** | 13(72) | 2(11) | 17(55) | 8(26) | 0.36  OR 0.47  (0.10-1.88) | 0.29  OR 2.73  (0.45-29.73) | 0.67 NS  OR 0.50  (0.06-3.45) | **<0.001**  **OR 0.17**  **(0.06-0.43)** |
| C/W WTA | 4(29) | 8(57) | 39(80) | 33(67) | <0.001  OR 0.11  (0.02-0.47) | 0.53  OR 0.65  (0.16-2.69) | 15(83) | 13(72) | 24(77) | 20(65) | 0.73 NS  OR 0.69  (0.10-3.64) | 0.75  OR 0.70  (0.15-2.87) | 0.25 NS  OR 3.18  (0.55-21.66) | 0.25 NS  OR 0.53  (0.19-1.44) |
| Table continues next page | | | | | | | | | | | | | | |
| Scenario 5 - | Grade IV SI with grade II liver laceration. HD stable on admission.  CT abdomen - Moderate IP blood. Contrast blush.  ATOMAC = NOM/TRANSFER. WTA = AE/TRANSFER. | | | | | | | | | | | | | |
| number | PS N=14(%) | | AS N=48(%) | | p – PS vs AS, OR (95%CI) | | M-AS N=18(%) | | R-AS N=30(%) | | p – M-AS vs R-AS OR (95%CI) | | p - PS 10 vs 18 | p - AS 10 vs 18 |
| age | 10 | 18 | 10 | 18 | 10 | 18 | 10 | 18 | 10 | 18 | 10 | 18 |  |  |
| OM | 1(7) | 1(7) |  | 2(4) |  |  |  |  |  | 2(7) |  |  | 0.83 NS | **<0.001** |
| NOM | 9(64) | 4(29) | 3(6) | 5(10) | 0.35 NS | 0.12 NS | 1(6) | 1(6) | 2(7) | 4(13) | 0.74 NS | 0.11 NS |  |  |
| AE | 3(21) | 5(36) | 13(27) | 31(65) |  |  | 4(22) | 15(83) | 9(30) | 16(53) |  |  |  |  |
| TF | 1(7) | 4(29) | 33(69) | 10(21) |  |  | 13(72) | 2(11) | 19(63) | 8(27) |  |  |  |  |
| C/W ATOMAC | 10(71) | 8(57) | 35(73) | 15(31) | 1 NS | 0.12 NS  OR 2.88  (0.73-12.07) | 14(78) | 3(17) | 21(70) | 12(40) | 0.74 NS  OR 0.67  (0.12-3.03) | 0.12 NS  OR 3.25  (0.69-21.31) | 0.69 NS  OR 0.55  (0.08-3.28) | **<0.001**  **OR 0.17**  **(0.06-0.44)** |
| C/W WTA | 4(29) | 9(64) | 45(94) | 41(85) | **<0.001**  **OR 0.03**  **(0.004-0.17)** | 0.12 NS  OR 0.31  (0.07-1.55) | 17(94) | 17(94) | 28(93) | 24(80) | 1  OR 0.82  (0.01-17.03) | 0.23 NS  OR 0.24  (0.005-2.28) | 0.12 NS  OR 4.24  (0.72-30.00) | 0.31 NS  OR 0.39  (0.06-1.87) |
| Scenario 6 - | Grade IV SI with grade II liver laceration. Surgeon suspects IA bleeding.  Initial response to 10mls/kg NS + 10mls/kg PRBCs (age 10)/NS bolus +1 Unit PRBCs (age 18).  Recurrent HD instability at 4 hours responsive to further blood products.  CT abdomen - Moderate IP blood. Contrast blush.  ATOMAC = consider AE/TRANSFER. WTA = OM/AE/TRANSFER. | | | | | | | | | | | | | |
| number | PS N=14(%) | | AS N=48(%) | | p – PS vs AS, OR (95%CI) | | M-AS N=18(%) | | R-AS N=30(%) | | p – M-AS vs R-AS OR (95%CI) | | p - PS 10 vs 18 | p - AS 10 vs 18 |
| age | 10 | 18 | 10 | 18 | 10 | 18 | 10 | 18 | 10 | 18 | 10 | 18 |  |  |
| OM | 1(7) | 2(14) | 17(35) | 26(54) |  |  | 4(22) | 10(56) | 13(43) | 16(53) |  |  | 0.74 NS | **<0.001** |
| NOM | 2(14) | 2(14) |  | 1(2) | **<0.001** | **<0.05** |  |  |  | 1(3) | 0.14 NS | 0.83 NS |  |  |
| AE | 10(71) | 8(57) | 8(17) | 17(35) |  |  | 2(11) | 7(39) | 6(20) | 10(33) |  |  |  |  |
| TF | 1(7) | 2(14) | 23(48) | 4(8) |  |  | 12(67) | 1(6) | 11(37) | 3(10) |  |  |  |  |
| C/W ATOMAC | 13(93) | 12(86) | 31(65) | 22(46) | **<0.05**  **OR 6.96**  **(0.89-319.98)** | **<0.05**  **OR 6.89**  **(1.31-70.07)** | 14(78) | 8(44) | 17(57) | 14(47) | 0.21  OR 0.38  (0.07-1.62) | 1 NS  OR 1.09  (0.29-4.18) | 1 NS  OR 0.47  (0.01-10.22) | 0.10 NS  OR 0.47  (0.19-1.14) |
| C/W WTA | 12(86) | 12(86) | 48(100) | 47(98) | **<0.05**  **OR 0**  **(0.00-1.50)** | 0.12 NS  OR 0.13  (0.002-2.76) | 18(100) | 18(100) | 30(100) | 29(97) | Uniform | 1  OR 0  (0-64.93) | 1 NS  OR 1  (0.06-15.95) | 1 NS  OR 0  (0.00-39.00) |
| Table continues next page | | | | | | | | | | | | | | |

| Scenario 9 | Grade IV SI. Intubated and ventilated. Surgeon suspects IA bleeding. Small extradural haematoma on head CT.  Initial response to 10mls/kg NS + 10mls/kg PRBCs (age 10)/NS bolus +1 Unit PRBCs (age 18).  Recurrent HD instability at 4 hours responsive to further blood products.  CT abdomen - Moderate IP blood. Contrast blush.  ATOMAC – consider AE/TRANSFER. WTA – AE/OM/TRANSFER. | | | | | | | | | | | | | |
| --- | --- | --- | --- | --- | --- | --- | --- | --- | --- | --- | --- | --- | --- | --- |
| number | PS N=14(%) | | AS N=47(%) | | p – PS vs AS, OR (95%CI) | | M-AS N=18(%) | | R-AS N=29(%) | | p – M-AS vs R-AS OR (95%CI) | | p - PS 10 vs 18 | p - AS 10 vs 18 |
| age | 10 | 18 | 10 | 18 | 10 | 18 | 10 | 18 | 10 | 18 | 10 | 18 |  |  |
| OM | 1(7) | 2(14) | 16(34) | 22(47) |  |  | 3(17) | 8(44) | 13(45) | 14(48) | **<0.05** | 0.79 NS | 0.40 NS | **<0.05** |
| NOM | 2(14) | 2(14) | 1(2) | 1(2) | **<0.001** | 0.09 NS | 1(6) | 1(6) |  |  |  |  |  |  |
| AE | 10(71) | 6(43) | 6(13) | 13(28) |  |  | 1(6) | 6(33) | 5(17) | 7(24) |  |  |  |  |
| TF | 1(7) | 4(29) | 24(51) | 11(23) |  |  | 13(72) | 3(17) | 6(21) | 3(10) |  |  |  |  |
| C/W ATOMAC | 13(93) | 12(86) | 31(66) | 25(53) | 0.08  OR 6.55  (0.84-302.22) | **<0.05**  **OR 5.15**  **(0.98-52.42)** | 15(83) | 10(56) | 16(55) | 15(52) | 0.06  OR 0.25  (0.04-1.18) | 1 NS | 1 NS  OR 0.47  (0.007-10.22) | 0.29 NS  OR 0.59  (0.23-1.46) |
| C/W WTA | 12(86) | 12(86) | 46(97) | 46(97) | 0.14  OR 0.14  (0.002-2.82) | 0.13 NS  OR 0.14  (0.002-2.82) | 17(94) | 17(94) | 29(100) | 29(100) | 0.38 | 0.38 | 1 NS  OR 1  (0.06-15.4) | 1 NS  OR 1  (0.01-80.13) |

| Supplementary Table 3e: Responses for scenario 10 by hospital category  Grade IV splenic injury with moderate intraperitoneal blood and no contrast blush on abdominal CT-scan. Pediatric guidelines: ATOMAC = the North American Paediatric Trauma Consortium (Arizona-Texas-Oklahoma-Memphis-Arkansas). Adult guidelines: WTA = Western Trauma Association. SI = splenic injury. HD=haemodynamic. IA = intraabdominal. IP = intra-peritoneal. NS=normal saline, PRBC=packed red blood cells. OM = operative management. NOM = non-operative management. AE = angioembolisation. PTC = paediatric trauma centre. ATC = metropolitan adult trauma centre. RTC = regional/rural trauma centre. MLH = metropolitan local hospital. RLH = regional/rural local hospital. OR = odds ratio. Unif = uniform. Transfer and NOM analysed as one variable for treatment choice and alignment with guidelines. * denotes each Paediatric Surgeon at an adult centre | | | | | | | | | | | | |
| --- | --- | --- | --- | --- | --- | --- | --- | --- | --- | --- | --- | --- |
| Scenario 10 | Grade IV SI. Intubated and ventilated. Surgeon suspects IA bleeding. Small extradural haematoma on head CT.  Initial response to 10mls/kg NS + 10mls/kg PRBCs (age 10)/NS bolus +1 Unit PRBCs (age 18).  Recurrent HD instability at 4 hours responds transiently to further blood products.  HD instability returns over next 2 hours with further appropriate resuscitation.  CT abdomen – Moderate IP blood. No contrast blush.  ATOMAC – consider NOM/AE/OM/TRANSFER. WTA – OM/TRANSFER. | | | | | | | | | | | |
| number | PTC N=12(%) | | ATC N=14(%) | | RTC N=20(%)* | | MLH N=5(%)* | | RLH N=9(%) | | p | |
| age | 10 | 18 | 10 | 18 | 10 | 18 | 10 | 18 | 10 | 18 | 10 | 18 |
| OM | 4(33) | 4(33) | 5(36) | 12(86) | 11*(55) | 12(60) | 2(40) | 3(60) | 4(44) | 5(55) |  |  |
| NOM | 3(25) | 3(25) |  |  |  | 1(5) |  |  | 1(11) | 1(11) | 0.27 NS | 0.07 NS |
| AE | 5(42) | 3(25) | 1(7) | 2(14) | 2(10) | 3!15) | 1*(20) |  |  |  |  |  |
| TF |  | 2(17) | 8(57) |  | 7(35) | 4*(20) | 2(40) | 2*(40) | 4(44) | 3(33) |  |  |
| C/W ATOMAC | 12(100) | 12(100) | 14(100) | 14(100) | 20*(100) | 20*(100) | 5*(100) | 5*(100) | 9(100) | 9(100) | uniform | uniform |
| C/W WTA | 4(33) | 6(50) | 13(93) | 12(86) | 18(90) | 16(80) | 4(80) | 5(100) | 8(89) | 8(89) | **<0.01** | 0.14 NS |

| Supplementary Table 3f: Responses for scenario 10 by surgeon type  Grade IV splenic injury with moderate intraperitoneal blood and no contrast blush on abdominal CT-scan. Pediatric Guidelines: ATOMAC = the North American Paediatric Trauma Consortium (Arizona-Texas-Oklahoma-Memphis-Arkansas). Adult guidelines: WTA = Western Trauma Association. SI = splenic injury. HD=haemodynamic. IA = intraabdominal. IP = intra-peritoneal. NS=normal saline, PRBC=packed red blood cells. OM = operative management. NOM = non-operative management. AE = angioembolisation. AS =adult surgeon. PS = paediatric surgeon. M = metropolitan. R = regional/rural. OR = odds ratio. Unif = uniform. Transfer and NOM analysed as one variable for treatment choice and alignment with guidelines. | | | | | | | | | | | | | | |
| --- | --- | --- | --- | --- | --- | --- | --- | --- | --- | --- | --- | --- | --- | --- |
| Scenario 10 | Grade IV SI. Intubated and ventilated. Surgeon suspects IA bleeding. Small extradural haematoma on head CT.  Initial response to 10mls/kg NS + 10mls/kg PRBCs (age 10)/NS bolus +1 Unit PRBCs (age 18).  Recurrent HD instability at 4 hours responds transiently to further blood products.  HD instability returns over next 2 hours with further appropriate resuscitation.  CT abdomen – Moderate IP blood. No contrast blush.  ATOMAC – consider NOM/AE/OM/TRANSFER. WTA – OM/TRANSFER. | | | | | | | | | | | | | |
| number | PS N=14(%) | | AS N=46(%) | | p – PS vs AS, OR (95%CI) | | M-AS N=18(%) | | R-AS N=28(%) | | p – M-AS vs R-AS OR (95%CI) | | p - PS 10 vs 18 | p - AS 10 vs 18 |
| age | 10 | 18 | 10 | 18 | 10 | 18 | 10 | 18 | 10 | 18 | 10 | 18 |  |  |
| OM | 5(36) | 4(29) | 21(46) | 32(70) |  |  | 7(39) | 15(83) | 14(50) | 17(61) | 0.79 NS | 0.17 NS | 0.32 NS | **<0.05** |
| NOM | 3(21) | 3(21) | 1(2) | 2(4) | **<0.01** | **<0.05** |  |  | 1(4) | 2(7) |  |  |  |  |
| AE | 6(43) | 3(21) | 3(6) | 5(11) |  |  | 1(6) | 2(11) | 2(7) | 3(11) |  |  |  |  |
| TF |  | 4(29) | 21(46) | 7(15) |  |  | 10(56) | 1(6) | 11(39) | 6(21) |  |  |  |  |
| C/W ATOMAC | 14(100) | 14(100) | 46(100) | 46(100) | uniform | uniform | 18(100) | 18(100) | 28(100) | 18(100) | uniform | uniform | uniform | uniform |
| C/W WTA | 5(36) | 8(57) | 42(91) | 39(85) | **<0.001**  **OR 0.06**  **(0.009-0.29**) | 0.06 NS  OR 024  (0.05-1.14) | 17(94) | 16(89) | 25(89) | 23(82) | 1 NS  OR 0.50  (0.009-6.81) | 0.69  OR 0.58  (0.05-4.12) | 0.45 NS  OR 2.32  (0.42-14.36) | 0.52 NS  OR 0.53  (0.11-2.29) |
